# The countdown paradox: time-varying analysis of biomarker-clock age and symptom onset

**DOI:** 10.64898/2026.06.19.26355994

**Authors:** Yuxin Zhu, Corinne Pettigrew, Anja Soldan, Abhay Moghekar, Marilyn Albert, Karen Bandeen-Roche, Mei-Cheng Wang

**Affiliations:** Department of Neurology, Johns Hopkins University School of Medicine, Baltimore, MD, USA; Department of Biostatistics, Johns Hopkins Bloomberg School of Public Health, Baltimore, MD, USA; Johns Hopkins Armstrong Institute for Patient Safety and Quality, Baltimore, MD, USA; Johns Hopkins Data Science and AI Institute, Baltimore, MD, USA

**Author notes:** Correspondence to: Yuxin Zhu, Department of Neurology, Johns Hopkins University School of Medicine, Baltimore, MD, USA.

**Keywords:** Alzheimer’s disease, mild cognitive impairment, survival analysis, biomarkers, biomarker clock

## Abstract

Research focused on Alzheimer’s disease (AD) ‘biomarker clocks’ seeks to identify ages at which AD pathological landmarks occur (e.g., initiation of amyloid accumulation) and are meaningfully related to disease outcomes (e.g., symptom onset). However, the statistical approach for assessing the association between age at biomarker-clock event and remaining time to clinical symptom onset can create a structural artifact. We term it here the ‘countdown paradox’, because the remaining time to symptom onset shrinks as the age at biomarker-clock event increases, which may result in inaccurate associations between age at biomarker-clock event and the remaining time.

We conducted analyses to examine this issue with simulation studies and theoretical results, and also examined it empirically using five biomarkers in two longitudinal AD-related cohorts (BIOCARD and ADNI): (1) CSF Aβ42/Aβ40, (2) CSF p-tau181, (3) plasma p-tau181, (4) amyloid PET, and (5) plasma p-tau217. As an alternative analytic approach to the standard approach, we used a time-varying effect analysis that evaluates the association between biomarker-clock events and symptom onset on the ‘age’ time scale, avoiding the structural coupling between predictor and outcome. This analytic approach generates clinically relevant insights on the prognostic value of biomarker-clock events.

Under simulated null scenarios in which the biomarker was generated independent of symptom onset, the standard analysis produced false-positive rates up to 100% and hazard ratios above 1, regardless of the true effect direction, whereas the time-varying analysis maintained type I error near the nominal 5%. Moreover, in analyses of both the BIOCARD and ADNI cohorts, the standard analysis produced uniformly significant associations for ages at biomarker-clock events, based on all five biomarkers (hazard ratios 1.94–3.34, all P < 0.01), comparable to the pattern predicted by the countdown paradox and reported in the literature. The time-varying analysis showed a different pattern for the effect of age at biomarker-clock events: for all biomarkers investigated, a younger age at biomarker-clock events is associated with a higher hazard for symptom onset on the age scale, conveying the opposite prognostic message implied by the standard analysis.

These findings suggest that the standard biomarker-clock analysis may generate inaccurate associations and even reverse the apparent direction of the age effect, inverting the resulting prognostic message. A time-varying effect analysis avoids this by relating the age at a biomarker-clock event to clinical onset, with important implications for interpreting prior biomarker-clock studies.

## 1. Introduction

Alzheimer’s disease (AD) neuropathological changes begin decades before clinical symptoms are observed^1–3^, including the accumulation of amyloid-beta plaques and tau tangle formation. These pathological brain changes can be estimated with biofluid and neuroimaging biomarkers. A better understanding of individualized patterns of biomarker change is crucial for early detection and risk stratification as treatments move to earlier disease stages. Within this context, AD ‘biomarker clocks’ have been proposed that estimate the age at which biomarker changes begin during the preclinical disease phase, with the goal of determining whether and how these estimated ages relate to meaningful disease outcomes, such as the onset of AD symptoms. These AD biomarker clocks calculate the age at which biomarker change begins or accelerates as an estimate of pathological landmarks^4–10^. For example, by fitting trajectory models to longitudinal biomarker measurements, researchers have estimated when amyloid or tau accumulation began for each person, anchoring the disease timeline to a biomarker-based clock rather than to clinical symptoms. Efforts to derive ages at biomarker-clock events (AABCs) have applied various approaches for biomarker trajectory modeling, pathological landmark event definition, and computing algorithms, and have been applied to a variety of biofluid and neuroimaging biomarkers across studies including the Alzheimer’s Disease Neuroimaging Initiative (ADNI)^11^, Biomarkers of Cognitive Decline Among Normal Individuals (BIOCARD)^12^, the Wisconsin Registry for Alzheimer’s Prevention^8^, and the Mayo Clinic Study of Aging^10^. It is critical to use valid analytic approaches to evaluate whether derived AABCs are associated with clinical symptom onset and to distinguish informative AABCs from non-informative ones. Informative AABCs can then be used as a basis for individualized prognosis, risk stratification, and identification of treatment windows for disease-modifying therapies.

In the existing literature, the standard analytical approach for evaluating the association of derived AABCs with AD clinical symptom onset regresses the remaining time from AABC to symptom onset on the AABC^4–6^, often applying the Cox proportional hazards models. These models produce hazard ratios associated with each variable included in the model; a hazard ratio significantly greater than 1 suggests that larger variable values are associated with shorter remaining time to onset, whereas a hazard ratio significantly less than 1 suggests that larger variable values are associated with longer remaining time to onset. In this paper, however, we describe how this standard approach can create a structural artifact.

As an intuitive example, consider a familiar quantity in longitudinal AD cohort studies: a participant’s age at study entry. When the remaining time from study entry to symptom onset is regressed on age at entry, a negative association tends to emerge because a later age at entry necessarily leaves less remaining time to symptom onset, given the natural lifespan limits.

Investigators routinely model the time from baseline to symptom onset while adjusting for baseline age this way and typically find an effect of the baseline age. Yet, investigators do not interpret the effect to mean that entering the study earlier is protective or that later enrollment accelerates symptom onset. Nor would they use study entry as a landmark event for disease prognosis in clinical settings, or advocate for early study entry based on these results. The negative association found in such analyses reflects how the remaining time is constructed, and a comparable effect would arise for other age-at-event variables, including ones for events with little or no relationship to disease.

The standard biomarker-clock analysis adopts the same remaining-time regression, on AABC instead of age at study entry, but results from such analyses have invited the very interpretations we would withhold for study entry age—that a later AABC is associated with higher risk and accelerates progression, and that the biomarker-clock event should be used for disease prognosis. **The concern lies not in the estimated effect itself but in the interpretation and the conclusion implied.** As we illustrate below, an AABC derived from a biomarker unrelated to the disease can still produce a hazard ratio above 1, often with a significant P value, which may be misinterpreted as indicating that a later AABC carries higher risk. Because this structural artifact can be present even for non-informative biomarkers, it can inflate the type I error (i.e., false positive rate) of the standard analysis. We term this the **countdown paradox**: the remaining time, conceptualized as the ‘countdown’ from the AABC to age at symptom onset, mechanically shrinks as the AABC increases. Figure 1 illustrates this phenomenon in a hypothetical cohort in which the biomarker-clock event is unrelated to symptom onset: the remaining time mechanically shrinks as that age increases (Fig. 1A), so the standard statistical analysis produces a strongly significant association (Fig. 1B). An alternative time-varying analysis on the age time scale, described in detail below, shows the absence of association (Fig. 1C).

**Figure 1.**
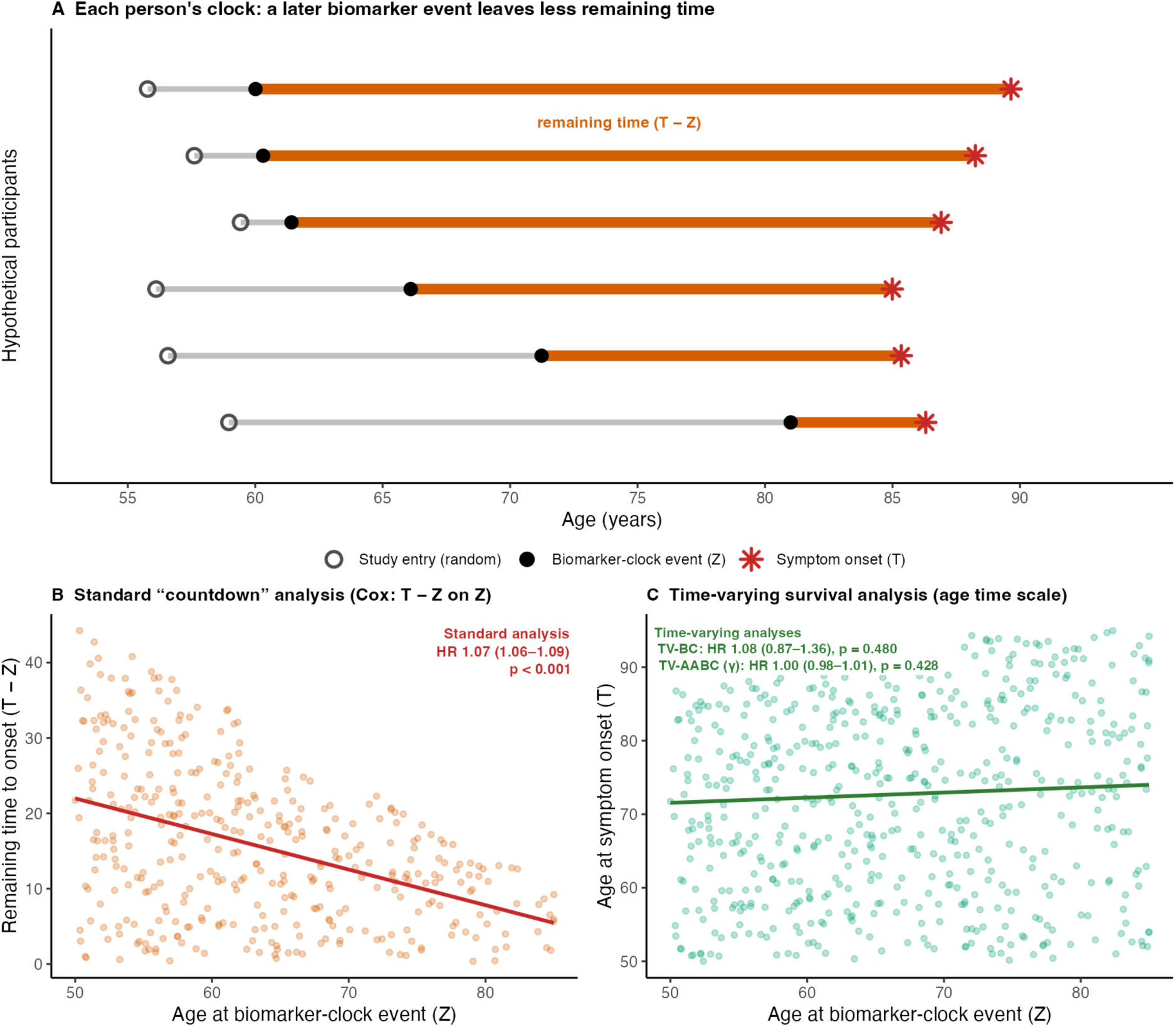
The countdown paradox, illustrated. (A) Age-scale timelines for six hypothetical participants. Each enters the study at a random age (open circle), becomes biomarker-positive at age Z (filled circle), and later develops symptoms at age T (asterisk); the remaining time from positivity to onset, T − Z (orange), mechanically shrinks as Z increases, even though the onset ages T are generated independently of Z. (B, C) A simulated cohort in which the age at the biomarker-clock event (Z) and the age at symptom onset (T) are drawn independently. (B) The standard ‘countdown’ analysis—a Cox model of the remaining time to onset (T − Z) on Z, among individuals whose onset is observed after the event—reports a strongly significant association (HR 1.07, 95% CI 1.06–1.09, p < 0.001) where none exists. (C) The same cohort analyzed on the age time scale: a time-varying biomarker-clock model (TV-BC: HR 1.08, 95% CI 0.87–1.36, p = 0.48) and a time-varying model with an age-at-positivity interaction (TV-AABC, γ: HR 1.00, 95% CI 0.98–1.01, p = 0.43) both correctly find no association. Scatterplots and trend lines are shown for illustration; the quoted hazard ratios are from the survival models, underscoring that relating the age at a biomarker-clock event to onset requires appropriate time-to-event methods rather than a remaining-time regression. Alt text: A three-part schematic. A timeline panel shows six participants on an age axis, each becoming biomarker-positive and later developing symptoms, with the gap between the two shrinking as the age at positivity increases even though onset ages are independent of it. A scatterplot shows the standard remaining-time analysis producing a steep, statistically significant slope, while a third panel shows the same data on the age time scale, where the time-varying analyses find a flat, non-significant relationship.

This structural artifact can be explained by exploring the underlying statistical structure. The outcome of remaining time from AABC to symptom onset is equal to age at symptom onset minus AABC, creating a mechanical coupling between remaining time and AABC: Covariance (remaining time, AABC) = Covariance (age at symptom onset, AABC) – Variance (AABC).

Under the null scenario where the biomarker is unrelated to disease, the first term of the formula vanishes, and the covariance reduces to negative Variance (AABC) (see Supplementary Note 1 for a detailed derivation). This guaranteed negative covariance produces a component of spurious association in regression analysis of remaining time on AABC. Consequently, this regression does not isolate the quantity of scientific interest, because the coupling muddles what is being estimated. Beyond a single biomarker, this artifact can propagate to analyses involving multiple biomarker-clock events. When the ages at successive events, for example, amyloid and tau positivity, are not appropriately adjusted for, the induced correlations can generate spurious associations among the intervals separating these events and symptom onset^6^. The presence of this artifact also explains the two seemingly contradictory findings in the literature: that cognitive decline is faster among individuals with younger AABCs (longer chronicity) ^5, 8, 12^, but that remaining time to symptom onset is shorter^4, 5^.

In this study, we first present simulation studies that characterize the mechanism and magnitude of the countdown paradox artifact. Then, we present results for five biomarkers across two cohorts: three biomarkers based on data from the BIOCARD cohort (CSF Aβ42/Aβ40, CSF p-tau181, and plasma p-tau181) and two biomarkers based on data from the ADNI cohort (amyloid PET and plasma p-tau217), using both the standard ‘countdown’ approach and the time-varying effect (TV) analytic approach that we propose. Taken together, the work aims to provide practical guidance for studying the effects of reaching biomarker-clock events and of AABCs on disease outcomes. Although we demonstrate the countdown paradox in the context of AD biomarkers, the phenomenon is not specific to any biomarker, disease, or computational algorithm used to derive biomarker positivity age. The proposed TV analytic approach can be applied to analogous biomarker-clock work in other neurodegenerative disorders, including that in Huntington’s disease^13, 14^, Parkinson’s disease, and dementia with Lewy bodies^15^. Although we focus on the standard analytic approach evaluated here, Cox proportional hazards model, models other than the Cox proportional hazards model are similarly predisposed to the countdown paradox, if they regress remaining time from AABC to symptom onset on AABC.

## 2. Materials and methods

### 2.1 Study populations

#### 2.1.1 BIOCARD cohort

The BIOCARD study is a prospective longitudinal cohort study that was conducted at the National Institutes of Health from 1995-2005 and designed to examine the earliest neurobiological features of AD^16^. Participants (n = 349) were cognitively unimpaired at enrollment, with a mean entry age of 56.8 ± 10.3 years (original cohort). The study was re-established at Johns Hopkins University in 2009, where annual clinical and cognitive assessments have continued. Serial cerebrospinal fluid and blood biomarker measurements were obtained approximately biennially from 1995–2005, and from 2010 to the present. Consensus clinical diagnoses of mild cognitive impairment (MCI) and dementia were made at each visit based on standard criteria, with the event time defined as the estimated age at MCI symptom onset, as described previously^16^. The present analysis included 238 participants for CSF biomarkers (83 MCI events; median follow-up 20.4 years) and 200 participants for plasma p-tau181 (37 MCI events; median follow-up 13.1 years).

#### 2.1.2 ADNI cohort

The Alzheimer’s Disease Neuroimaging Initiative (ADNI) is a multisite longitudinal study that began in 2004 (as ADNI-1) with subsequent phases (ADNI-GO, ADNI-2, ADNI-3, ADNI-4). The present analyses included participants who were cognitively unimpaired at the analytic baseline, with longitudinal clinical assessments and amyloid PET scans (florbetapir or florbetaben) or available plasma p-tau217 measurements. The clinical outcome was defined as the age at the first ADNI visit with a diagnosis of MCI or dementia (7/106 or 6.6% of progressors in the amyloid PET cohort, and 7/162 or 4.3% of progressors in the plasma p-tau217 cohort had a first impaired-diagnosis label of dementia without a preceding documented MCI visit; we refer to this outcome as ‘MCI onset’ for simplicity). The analysis included 575 participants for the amyloid PET analysis (106 MCI events; median follow-up 5.7 years) and 801 for the plasma p-tau217 analysis (162 MCI events; median follow-up 3.5 years).

### 2.2 Biomarker measurements and age at positivity

#### 2.2.1 AABC estimation

For each of the five biomarkers from two cohorts, we used the Sampled Iterative Local Approximation (SILA)^5^, which estimates a value-vs-time trajectory by modeling the rate of biomarker change as a function of biomarker value and integrating it over time, then aligns each participant’s measurements to that trajectory to estimate their AABC. We also refer to AABC as the age at biomarker positivity. Participants whose anchored trajectory does not cross the positivity threshold within the feasible age range are coded as having no valid AABC, and are excluded from the standard analysis but contribute not-yet-positive person-time to the time-varying analyses. Within each biomarker analysis, AABCs were standardized by z-scoring among participants who reached biomarker positivity, so HR-γ in the TV-AABC (time-varying age-at-biomarker-clock effect) model is interpreted as the hazard ratio associated with a one-standard-deviation increase in AABC. All analyses included participants who met the cohort inclusion criterion (i.e., cognitively unimpaired at the analytic baseline) with complete covariate data.

#### 2.2.2 BIOCARD CSF biomarkers

Cerebrospinal fluid biomarkers were measured at approximately biennial visits. Samples were collected by lumbar puncture and stored at −80 °C, and Aβ40, Aβ42, and p-tau181 were assayed on a fully automated electrochemiluminescence platform (Lumipulse G1200; Fujirebio Diagnostics, Inc.). Two CSF biomarkers were analyzed: the Aβ42/Aβ40 ratio and phosphorylated tau 181 (p-tau181)^17^. Positivity thresholds were ≤0.085 for the CSF Aβ42/Aβ40 ratio (lower values indicate amyloid accumulation) and ≥35 pg/mL for CSF p-tau181. Positivity thresholds for CSF Aβ42/Aβ40 and CSF p-tau181 were derived by examining the rate of biomarker change as a function of biomarker value and selecting the threshold beyond which the rate of change becomes non-zero in the worsening direction, consistent with the prior analysis on estimated age of CSF AD biomarker change^12^.

#### 2.2.3 BIOCARD Plasma p-tau181

Plasma p-tau181 was measured using the Quanterix Simoa platform^18, 19^. Although BIOCARD collected plasma during both an earlier NIH phase and the subsequent Johns Hopkins phase starting in 2010, we restricted the analysis to measurements over the latter phase to avoid confounding by differences in sample collection procedures between study phases. The TV analysis for plasma used the age at the first Johns Hopkins-phase plasma measurement as the left-truncation (analytic baseline) age and required participants to be cognitively unimpaired at that visit, to avoid overextrapolation of the SILA algorithm, reducing sample size and event count relative to the CSF analyses. The SILA-based trajectory modeling was used to estimate age at plasma p-tau181 positivity, using a threshold of ≥0.8 pg/mL, selected to yield a positivity rate around 50%, similar to CSF biomarkers in BIOCARD, given that the literature varies regarding an appropriate cutoff to use^20^.

#### 2.2.4 ADNI Amyloid PET

Amyloid PET imaging was performed using florbetapir (AV45) or florbetaben (FBB). Age at amyloid onset was estimated using SILA, with a positivity threshold of ≥1.11 SUVR (whole cerebellum reference) for florbetapir^21^.

#### 2.2.5 ADNI Plasma p-tau217

Plasma p-tau217 was measured using the Fujirebio Lumipulse platform. Age at positivity was estimated using SILA trajectory modeling applied to plasma measurements, using a positivity threshold of ≥0.300 pg/mL, the UPenn ADRC high-confidence-positive cutoff validated against florbetaben PET^22^.

### 2.3 Clinical outcome

The outcome was estimated age at onset of MCI for BIOCARD and age at MCI diagnosis for ADNI. In the ADNI analytic cohorts, 6.6% (7/106) of progressors in the amyloid PET analysis and 4.3% (7/162) in the plasma p-tau217 analysis received a dementia diagnosis without a documented preceding MCI diagnosis. For simplicity, we refer to this outcome as MCI onset. Participants without MCI onset were censored at the age of their last clinical assessment. Covariates for all analyses included sex, years of education (standardized), and APOE ε4 carrier status (*APOE* ε4 non-carriers = 0, *APOE* ε4 carriers = 1).

### 2.4 Statistical analysis

#### 2.4.1 Standard countdown analysis

The standard countdown analysis regressed remaining time from the biomarker-clock event to age at MCI onset on AABC using a Cox proportional hazards model, while adjusting for sex, education, and *APOE* ε4 carrier status. These models were restricted to participants whose estimated positivity age preceded MCI onset; this is the approach used in published biomarker-clock studies^4–6^. We apply this method in simulation analyses and analyses of BIOCARD and ADNI data to illustrate the countdown paradox and for comparison with TV analysis.

#### 2.4.2 Time-varying effect analysis

The time-varying (TV) effect analysis avoids the structural coupling between predictor and outcome by using age as the time scale, modeling biomarker positivity as a time-dependent indicator, and allowing AABC to have a time-varying effect. Each participant contributes person-time in two states: not-yet-positive before their estimated age at positivity, and positive after. Participants already positive at the analytic baseline contribute only positive person-time; those who never become positive contribute only not-yet-positive person-time. Data are structured in start-stop counting process format with left truncation at each participant’s analytic baseline, the age at which at-risk follow-up begins (the first study visit for the BIOCARD CSF and ADNI analyses, and the first Johns Hopkins-phase plasma measurement for BIOCARD plasma p-tau181), and right censoring at the last clinical visit if no MCI onset.

This approach is an established solution for immortal time bias in pharmacoepidemiology^23, 24^, a problem recognized in clinical domains^25–28^. It resolves two problems in the standard countdown analysis: (a) the immortal time component, because participants can contribute unexposed person-time before their positivity event; and (b) the structural covariance, by using the outcome of age at MCI onset instead of remaining time.

### TV-BC formulation

The TV-BC (time-varying biomarker-clock) model is a Cox proportional hazards model with age as the time scale while accommodating left truncation and right censoring, where the time-varying positivity indicator and covariates (sex, education, *APOE* ε4 carrier status) are included as predictors. This model investigates whether reaching biomarker positivity increases the hazard of MCI onset. A hazard ratio greater than 1 indicates biomarker-positive individuals have a higher hazard of MCI onset than not-yet-positive individuals.

### TV-AABC formulation

The TV-AABC model extends the TV-BC model by adding an interaction between the time-varying biomarker positivity indicator and the age at positivity among positive participants, with the same covariates for adjustment. In this model, HR-β represents the hazard of MCI onset for biomarker-positive individuals (positivity at AABC centering age) versus not-yet-positive individuals, while HR-γ captures how AABC affects the hazard among positive individuals, which is the question the standard countdown analysis attempts to address but cannot answer without incurring the structural artifact. Age at positivity was z-scored within each biomarker analysis among participants who have reached biomarker positivity. HR-γ is therefore interpreted as the hazard ratio associated with a one-standard-deviation increase in age at biomarker positivity.

#### 2.4.3 Simulation study design

### Study 1: Direct generation

Study 1 used 15 scenarios to isolate the structural mechanism of the countdown paradox under straightforward data-generating processes. In each scenario, the age at biomarker-clock event and the age at MCI onset were generated from independent distributions spanning uniform, normal, Weibull, gamma, beta, and exponential families. This construction yielded two variables unrelated by design, so any apparent association from the standard countdown analysis must reflect the structural artifact rather than biology or estimation noise. Seven of these scenarios varied the shape and parameterization of the distributions used to generate the age at biomarker-clock event and onset, together with an empirically calibrated setting anchored to the BIOCARD cohort, while holding sample size fixed at n = 500. Six additional sample sizes (n = 150 to 1,000) under two representative scenarios examined whether the bias diminishes as sample size grows. Two further scenarios, reported in Supplementary Note 2, examined sensitivity to the degree of right-censoring. Both the standard countdown analysis and the time-varying analyses were applied to every simulated dataset. All simulations used 1,000 replications per scenario. Per-scenario specifications and full results are provided in Supplementary Table 1.

### Study 2: LMM-based trajectory generation

Study 2 used 59 scenarios to evaluate the countdown paradox under realistic longitudinal biomarker dynamics. Biomarker measurements were simulated from a subject-specific linear mixed-effects model capturing both baseline level and trajectory slope, and ages at MCI onset were drawn conditionally on each participant’s baseline biomarker value through a proportional-hazards model with constant baseline hazard. Study baseline age was drawn from a uniform distribution over the scenario-specified entry-age window. Of the 59 scenarios, 19 were null scenarios in which the biomarker had no effect on the hazard of onset, used to characterize Type I error; the remaining 40 were non-null scenarios spanning true effect sizes from strongly protective to strongly harmful, used to characterize power and direction accuracy. Scenarios were organized along five design features, each varied across a range of values while others were held fixed at a reference setting: sample size, between-subject trajectory heterogeneity, baseline hazard of MCI onset, measurement precision of the biomarker, and cohort observation profile—that is, the entry-age window, visit cadence, maximum follow-up length, and reference baseline hazard—evaluated at two presets spanning contrasting study designs. These single-factor variations were complemented by a set of scenarios in which multiple design features were varied simultaneously. All simulations used 1,000 replications per scenario. Per-scenario specifications and full results across the 59 scenarios are provided in Supplementary Tables 2–7, organized by sensitivity category.

### Metrics

Primary metrics for summarizing simulation results included rejection rate (representing type I error under the null and power under the alternative), mean hazard ratio, and direction accuracy (proportion of replications with the hazard ratio in the correct direction).

### 2.5 Ethics

This study is a secondary analysis of de-identified data from two studies of human participants and did not involve collecting new data. The BIOCARD study was approved by the Institutional Review Board at Johns Hopkins, and all BIOCARD participants provided written informed consent. The Alzheimer’s Disease Neuroimaging Initiative (ADNI) was approved by the institutional review boards of all participating institutions, and written informed consent was obtained from all ADNI participants or their authorized representatives.

## 3. Results

### 3.1 Simulation demonstration

We first used simulation studies to demonstrate the countdown paradox under conditions where the truth is known (full simulation design in Methods; complete results in Supplementary Notes 3–4, Supplementary Tables 1–7). Under null scenarios where the biomarker was generated independently of symptom onset, the standard countdown analysis produced false positive rates of 100% across all scenarios in Study 1 (partial results in Fig. 2A; see Supplementary Table 1 for full results). TV-BC maintained type I error around 5% (4.0–6.0%), as did TV-AABC HR-β (3.8–5.6%) and TV-AABC HR-γ (4.4–5.5%; Fig. 2B and 2C). Under realistic biomarker trajectory scenarios (Study 2), the standard Cox model analysis showed inflated type I error ranging from 30.6% to 100% across scenarios (Fig. 2D) and frequently identified the wrong effect direction for harmful biomarkers (a biomarker is considered harmful if more abnormal values are associated with increased hazard of symptom onset; Fig. 2E). The TV approaches, in contrast, tracked the true effect direction in 70.7–100.0% of replications across non-null scenarios. These results suggest significant associations found by the standard countdown analysis can be misleading: the method can reject the null hypothesis even when no true association exists and often misidentifies the direction of effects when true effects are present.

**Figure 2.**
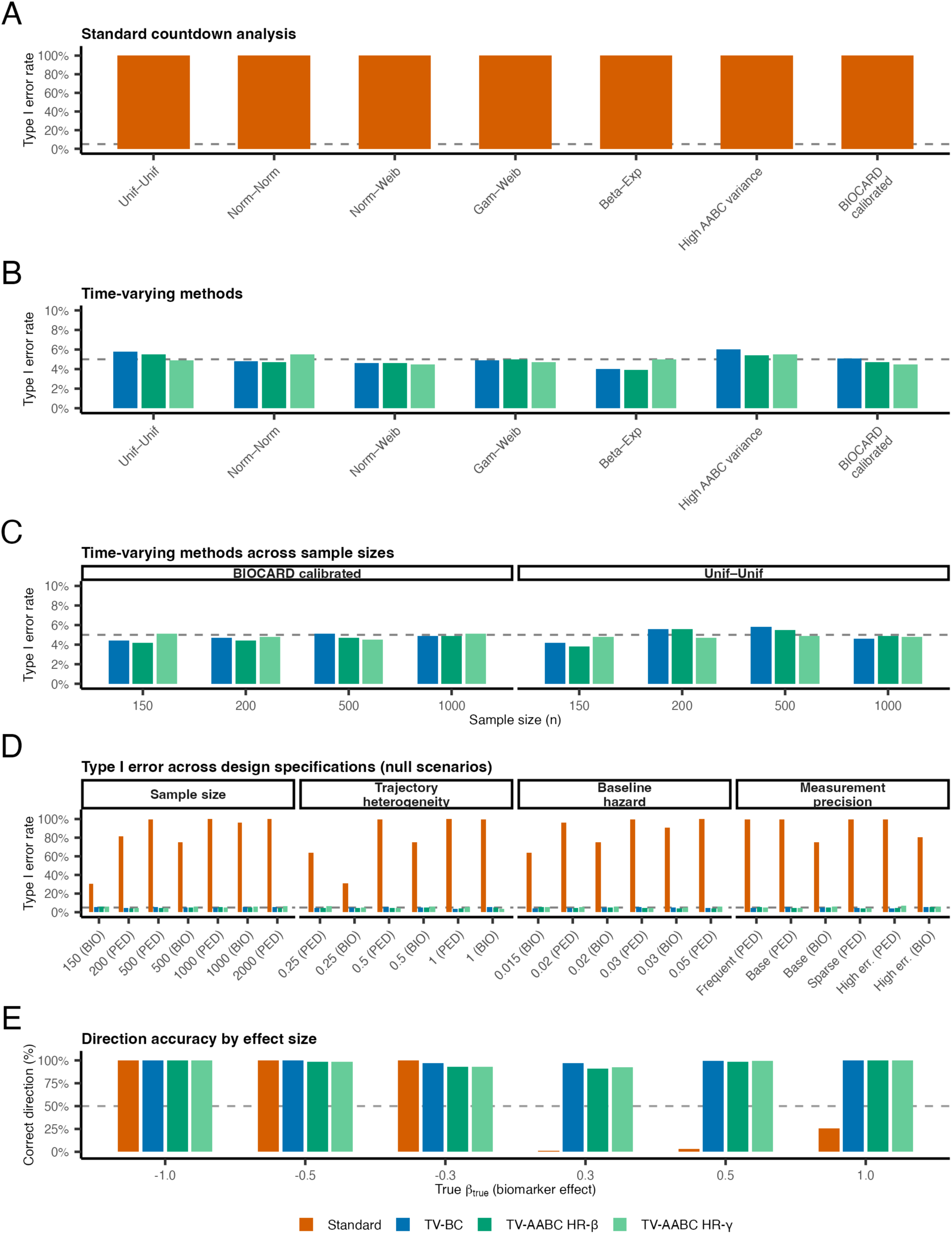
Simulation results from Study 1 (A–C) and Study 2 (D–E). Type I error and direction accuracy for four methods: Standard countdown (orange), TV-BC (blue), TV-AABC HR-β (blue green), and TV-AABC HR-γ (light green). The dashed line indicates the nominal 5% significance level. (A–C) Study 1 simulation results demonstrating the countdown paradox under direct distribution generation. (A) Type I error across 7 distribution-shape scenarios (S1–S5, S8, S9; all at n = 500). The standard countdown analysis produces false positive rates of 100% in every scenario. (B) TV method type I error at magnified scale (0–10%) for the same 7 scenarios, confirming rates near the nominal level for all three TV estimates. (C) Type I error across 8 sample-size scenarios (S1 and S9 at n = 150, 200, 500, 1,000). (D–E) Study 2 simulation results under realistic biomarker trajectories. (D) Type I error across 19 null scenarios spanning four design specification changes: sample size (n = 150–2,000), trajectory heterogeneity, baseline hazard, and measurement precision. Each specification was evaluated under two parameter regimes, pedagogical (PED) and BIOCARD-resembling (BIO), shown as separate bars on the x-axis. The standard countdown analysis shows inflated type I error (30.6–100%) across scenarios; TV methods maintain nominal rates. (E) Direction accuracy (proportion of replications identifying the correct effect direction) across true effect sizes. The standard countdown analysis produces HR > 1 in 74.5–100% of replications regardless of the true direction; TV-BC achieves 93.2–100.0% direction accuracy. See Supplementary Note 2 for scenario specifications. TV-AABC, time-varying age-at-biomarker-clock interaction model; TV-BC, time-varying biomarker-clock model. Alt text: A five-panel figure of bar charts (A–E); the y-axis is type I error rate (A–D) or correct-direction percentage (E), with a dashed reference line at 5% (A–D) or 50% (E). Panels A–C show Study 1: (A) the standard countdown analysis has type I error near 100% in all seven distribution scenarios; (B) the same seven scenarios on a magnified 0–10% scale, where the three time-varying estimates (TV-BC, TV-AABC HR-β, TV-AABC HR-γ) sit at or near the 5% line; (C) the time-varying methods stay near 5% across sample sizes of 150–1,000 for two representative scenarios. Panels D–E show Study 2 under realistic trajectories: (D) across null scenarios grouped into four design specifications (sample size, trajectory heterogeneity, baseline hazard, measurement precision) and two regimes (PED, BIO), the standard analysis is inflated by a scenario-dependent amount (roughly 30–100%) while the time-varying methods stay near zero; (E) grouped bars of the percentage of replications recovering the correct effect direction at six true effect sizes from −1 to +1; the standard analysis is high for negative (protective) effects but drops sharply toward zero for positive (harmful) effects, reflecting its bias toward a hazard ratio above 1, whereas the time-varying methods stay high across the range.

### 3.2 Analysis of BIOCARD data

For BIOCARD data (see Table 1 for demographic and clinical characteristics), SILA trajectory modeling was applied to 281 CSF and 203 plasma p-tau181 participants; of these, 131 (46.6%), 162 (57.7%), and 97 (47.8%) reached positivity by their last measurement for CSF Aβ42/Aβ40, CSF p-tau181, and plasma p-tau181, respectively, with the remaining participants’ trajectories not crossing the positivity threshold. After restricting to the TV analysis cohort with complete covariates (238 CSF participants with 83 MCI events; 200 plasma participants with 37 MCI events), 114, 148, and 97 had valid estimated AABCs [CSF Aβ42/Aβ40, mean (SD) 56.8 (9.5); CSF p-tau181, 59.3 (11.9); and plasma p-tau181, 66.0 (10.3) years, respectively].

**Table 1.** Participant characteristics.

| Characteristic | BIOCARD – CSF | BIOCARD – plasma | ADNI – Amyloid PET | ADNI – Plasma p-tau217 |
| --- | --- | --- | --- | --- |
| N | 238 | 200 | 575 | 801 |
| Progressors, n (%) | 83 (34.9%) | 37 (18.5%) | 106 (18.4%) | 162 (20.2%) |
| Female, n (%) | 146 (61.3%) | 125 (62.5%) | 333 (57.9%) | 482 (60.2%) |
| Years of education, mean (SD) | 17.1 (2.4) | 17.3 (2.2) | 16.5 (2.5) | 16.6 (2.5) |
| APOE e4 carrier, n (%) | 84 (35.3%) | 68 (34.0%) | 183 (31.8%) | 284 (35.5%) |
| Age at analytic baseline (in years), mean (SD) | 56.7 (10.3) | 64.5 (8.9) | 71.7 (6.8) | 70.2 (7.2) |
| Years of clinical follow-up since baseline, mean (SD) | 19.8 (5.7) | 11.9 (2.9) | 5.9 (4.4) | 4.3 (4.1) |

The standard countdown analysis found significant associations for all three biomarkers with remaining time to MCI onset (Table 3): CSF Aβ42/Aβ40 HR = 2.08 (95% CI 1.44–3.01, P < 0.001; n = 114, 54 events), CSF p-tau181 HR = 2.96 (95% CI 1.99–4.39, P < 0.001; n = 142, 56 events), and plasma p-tau181 HR = 2.34 (95% CI 1.39–3.94, P < 0.01; n = 95, 29 events). All hazard ratios exceeded 1.0, consistent with the countdown paradox prediction. These results reproduce the pattern reported in biomarker-clock studies that later positivity is associated with shorter remaining time to MCI onset^5^. However, this pattern should not be assumed to reflect a biologically meaningful association between AABC and hazard of MCI onset.

TV-BC found reaching biomarker positivity significantly increased the hazard of MCI onset for CSF Aβ42/Aβ40 (HR-β = 3.02, 1.80–5.05, P < 0.001) and plasma p-tau181 (HR-β = 3.28, 1.42–7.61, P < 0.01), with non-significant trends for CSF p-tau181 (HR-β = 1.52, 0.91–2.54, P = 0.11). TV-AABC showed concordant results: reaching biomarker positivity significantly increased the hazard of MCI onset for CSF Aβ42/Aβ40 (HR-β = 3.25, 1.93–5.48, P < 0.001) and plasma p-tau181 (HR-β = 2.74, 1.18–6.36, P < 0.05), but not CSF p-tau181 (HR-β = 1.45, 0.87–2.40, P = 0.15). Among positive individuals, earlier positivity (i.e., younger age at positivity) was associated with higher hazard of MCI onset for all three biomarkers: CSF Aβ42/Aβ40 HR-γ = 0.71 (0.51–0.99, P < 0.05), CSF p-tau181 HR-γ = 0.58 (0.42–0.78, P < 0.001), and plasma p-tau181 HR-γ = 0.51 (0.35–0.74, P < 0.001). The TV results contrast with the significant standard countdown results: biomarker positivity increases MCI risk (significant TV-BC for two of three biomarkers), and among positive individuals, earlier positivity is associated with higher hazard of MCI onset and younger MCI onset age, which may reflect more advanced pathology at younger ages. Full model coefficients are reported in Supplementary Table 8.

### 3.3 Analysis of ADNI data

For ADNI data, SILA trajectory modeling (trained on participants with ≥2 measurements) was applied to 1,373 participants with amyloid PET data and 1,546 with plasma p-tau217 data; of these, 741 (54.0%) and 512 (33.1%) reached positivity by their last measurement for amyloid PET and plasma p-tau217, respectively. After restricting to the TV analysis cohort with complete covariates (575 for amyloid PET with 106 MCI events; 801 for plasma p-tau217 with 162 events), 238 and 135 had valid estimated AABCs [mean (SD) 70.7 (8.7) and 75.3 (8.5) years, respectively]. Table 1 presents demographic and clinical characteristics.

For the amyloid PET analysis, the standard countdown analysis yielded a significant association between amyloid positivity and remaining time to MCI onset (HR = 3.34, 95% CI 2.41–4.61, P < 0.001; n = 236, 66 events), consistent with the countdown paradox prediction. TV-BC found a significant effect for reaching amyloid PET positivity: HR-β = 2.77 (1.84–4.17, P < 0.001), suggesting the presence of amyloid positivity increases the hazard of MCI onset. TV-AABC showed a significant effect of reaching amyloid positivity on MCI onset (HR-β = 2.89, 1.91–4.37, P < 0.001) but a non-significant effect of AABC (HR-γ = 0.87, 0.64–1.18, P = 0.36), indicating that while amyloid positivity increases the hazard of MCI onset, age at amyloid onset does not significantly impact the hazard among positive individuals in the ADNI cohort.

For the plasma p-tau217 analysis, the standard countdown analysis yielded a significant association with HR = 1.94 (95% CI 1.34–2.80, P < 0.001; n = 122, 48 events), again consistent with the countdown paradox prediction. TV-BC found a significant effect of reaching plasma p-tau217 positivity (HR-β = 2.60, 95% CI 1.83–3.69, P < 0.001), indicating plasma p-tau217 positivity increases the hazard of MCI onset. TV-AABC showed a significant effect of having experienced biomarker positivity (HR-β = 2.89, 95% CI 2.05–4.08, P < 0.001) and a significant effect of AABC (HR-γ = 0.46, 95% CI 0.32–0.66, P < 0.001), indicating that among positive individuals, earlier positivity is associated with higher hazard of MCI onset, consistent with results from the BIOCARD analyses.

### 3.4 Cross-cohort comparison

Across both cohorts, biomarker positivity is associated with a significantly increased hazard of MCI onset (HR-β) for four of five biomarkers under both the TV-BC and TV-AABC models; the exception is BIOCARD CSF p-tau181, for which the positivity effect is not significant (TV-BC HR-β = 1.52, P = 0.11; TV-AABC HR-β = 1.45, P = 0.15). The two amyloid measures, BIOCARD CSF Aβ42/Aβ40 (soluble amyloid) and ADNI amyloid PET (amyloid plaques), give concordant HR-β across modalities and cohorts. The cohorts differ in AABC modulation (HR-γ): BIOCARD shows a significant earlier-AABC effect on hazard across all three biomarkers, whereas ADNI shows this modulation only for plasma p-tau217 and not for amyloid PET (Table 3; Fig. 3).

**Figure 3.**
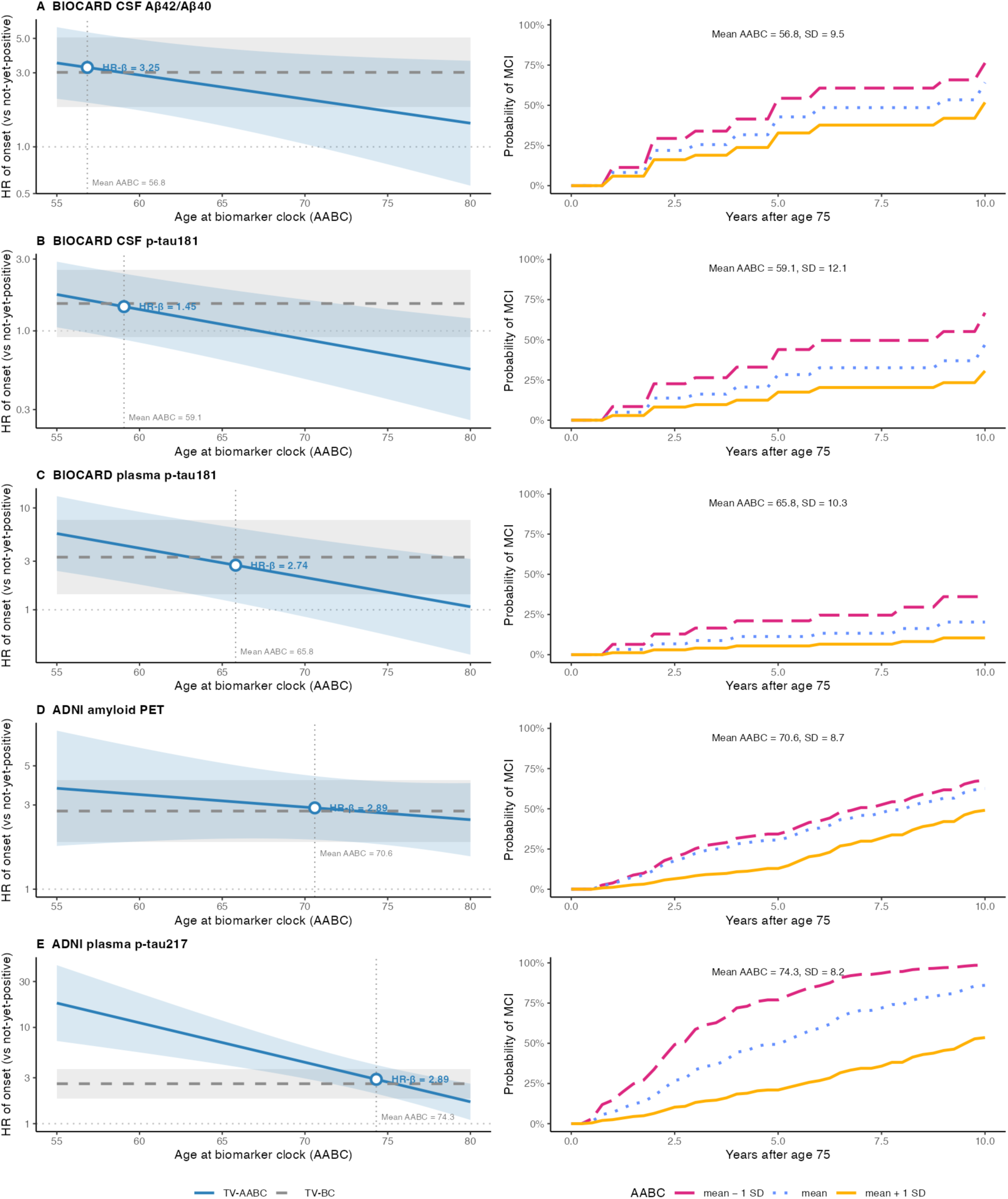
Hazard ratio of MCI onset and predicted onset risk given age at biomarker clock (AABC), across five biomarkers from two cohorts: (A) BIOCARD CSF Aβ42/Aβ40, (B) BIOCARD CSF p-tau181, (C) BIOCARD plasma p-tau181, (D) ADNI amyloid PET, (E) ADNI plasma p-tau217. Left column: hazard ratio (HR) of MCI onset as a function of AABC. Blue solid curve, TV-AABC interaction-model HR with 95% confidence ribbon; grey dashed line, TV-BC constant HR with light-grey 95% confidence interval. The vertical dotted line marks the mean AABC within each biomarker-positive subsample, and the open blue circle marks HR-β, the TV-AABC HR at the mean AABC (the model-centering value, the mean age at positivity among participants contributing observed biomarker-positive person-time, which can differ slightly from the descriptive mean AABC in Table 2). Across all five biomarkers the HR of MCI onset, relative to individuals without a biomarker-clock event, decreases as AABC increases; the shared 55–80-year x-axis covers all five mean AABCs and may imply extrapolation where those ages are absent from a given analysis. Right column: for the same TV-AABC model, the predicted probability of MCI onset within 10 years for an individual who is symptom-free at age 75, for AABC equal to the model-centering mean and to mean ± 1 SD (mean − 1 SD, pink long-dashed; mean, blue dotted; mean + 1 SD, yellow solid). Where AABC exceeds the landmark age of 75 the individual is not yet biomarker-positive at the landmark, so the curve follows the biomarker-negative baseline hazard until the age at positivity and the positive-state trajectory thereafter; other covariates are at the reference profile, and the y-axis is shared across panels. Per-biomarker HR-β and HR-γ (per 1 SD of AABC) are reported in Table 3; full model coefficients are in Supplementary Table 8. TV-AABC, time-varying age-at-biomarker-clock interaction model; TV-BC, time-varying biomarker-clock model. Alt text: A five-row, two-column figure, one row per biomarker. The left column plots the hazard ratio of symptom onset against age at biomarker positivity; the time-varying HR declines as age at biomarker positivity increases, shown against the TV-BC HR reference. The right column translates these into predicted onset probabilities over 10 years from age 75 for earlier, mean, and later ages at biomarker positivity.

**Table 2.** Degeneracy assessment by biomarker and cohort.

| | BIOCARD<br>CSF<br>Aβ42/Aβ40 | BIOCARD<br>CSF p-tau181 | BIOCARD<br>Plasma p-tau181 | ADNI<br>Amyloid PET | ADNI<br>Plasma p-tau217 |
| --- | --- | --- | --- | --- | --- |
| N with valid<br>SILA estimates | 114 | 148 | 97 | 238 | 135 |
| Already<br>positive at the<br>analytic<br>baseline, n (%) | 68 (59.6%) | 66 (44.6%) | 54 (55.7%) | 160 (67.2%) | 64 (47.4%) |
| Transition<br>during follow-up, n (%) | 46 (40.4%) | 76 (51.4%) | 41 (42.3%) | 76 (31.9%) | 58 (43.0%) |
| Never positive,<br>n | 124 | 96 | 105 | 339 | 679 |
| AABC, mean<br>(SD) year | 56.8 (9.5) | 59.3 (11.9) | 66.0 (10.3) | 70.7 (8.7) | 75.3 (8.5) |

Note: ‘N with valid SILA estimates’ counts all participants with a valid estimated age at biomarker positivity, including those whose estimated positivity age falls after their last clinical follow-up (extrapolated future positivity); these participants are also counted under ‘Never positive’, because their observed status remained negative throughout follow-up. The three observed-status categories—already positive at the analytic baseline, transition during follow-up, and never positive—partition the analysis cohort and sum to N. The AABC row reports the descriptive mean (SD) over these participants; Figure 3 instead uses the model-centering mean (over participants contributing biomarker-positive person-time), which differs materially only for ADNI plasma p-tau217 (74.3 vs 75.3 years).

This pattern may be explained by differences in effect size, in the cohorts, and in baseline age and the lengths of follow-up. BIOCARD CSF participants entered at mean age 56.7 with median 20.4 years of follow-up, and the plasma p-tau181 analysis entered at the Johns Hopkins–phase baseline at mean age 64.5 with median 13.1 years of follow-up; ADNI participants entered at mean age 71.7 with median 5.7 years of follow-up for amyloid PET, and mean age 70.2 with median 3.5 years of follow-up for plasma. Across BIOCARD biomarkers, 65–78% of MCI events occurred during biomarker-positive person-time, providing ample positive-state events for detecting how AABC affects risk (HR-γ). In ADNI, the pattern differed by biomarker. For amyloid PET, 62% of events occurred during biomarker-positive person-time, reflecting the high proportion positive at the analytic baseline (67.2%). Events in both states (40 pre-positive, 66 post-positive) provide adequate power for HR-β, while the non-significant HR-γ (0.87, P = 0.36) may reflect a smaller AABC effect size for amyloid PET. For plasma p-tau217, 70% of events occurred during biomarker-negative person-time. Despite fewer positive-state events (48), the significant HR-γ (0.46, P < 0.001) likely reflects a large effect size detectable with limited transitions.

Fig. 3 (left column) visualizes the TV-AABC interpretation by plotting the HR as a function of AABC for each biomarker and cohort, with TV-BC overlaid as a constant reference. The HR-β point at mean AABC corresponds to the effect reported in Table 3; the slope of the curve is governed by HR-γ. Across biomarkers, HR-γ ranges from nearly flat (ADNI amyloid PET, HR-γ = 0.87 per SD) to steep (ADNI plasma p-tau217, HR-γ = 0.46 per SD), indicating the prognostic significance of earlier versus later biomarker positivity differs across biomarkers. When HR-γ is close to 1, TV-BC tracks TV-AABC closely near the mean AABC, supporting TV-BC as a valid population-average summary; where HR-γ is far from 1, TV-AABC indicates further effects of AABC beyond TV-BC results. The right column of Fig. 3 translates these hazard ratios into absolute risk, showing the predicted probability of MCI onset over the decade following age 75, as an example, for individuals reaching positivity at the model-centering mean AABC and at one SD earlier or later. The spread among these curves mirrors HR-γ: for ADNI plasma p-tau217 the predicted 10-year probability rises from 54% with later positivity to 86% at the mean and 99% with earlier positivity, and for BIOCARD CSF Aβ42/Aβ40 from 52% to 64% to 76%, whereas for ADNI amyloid PET the three curves remain close (49–68%). These absolute risks make the prognostic consequences of earlier versus later positivity concrete: where AABC modulates hazard, crossing the biomarker threshold earlier corresponds to a higher risk of symptom onset.

**Table 3.**
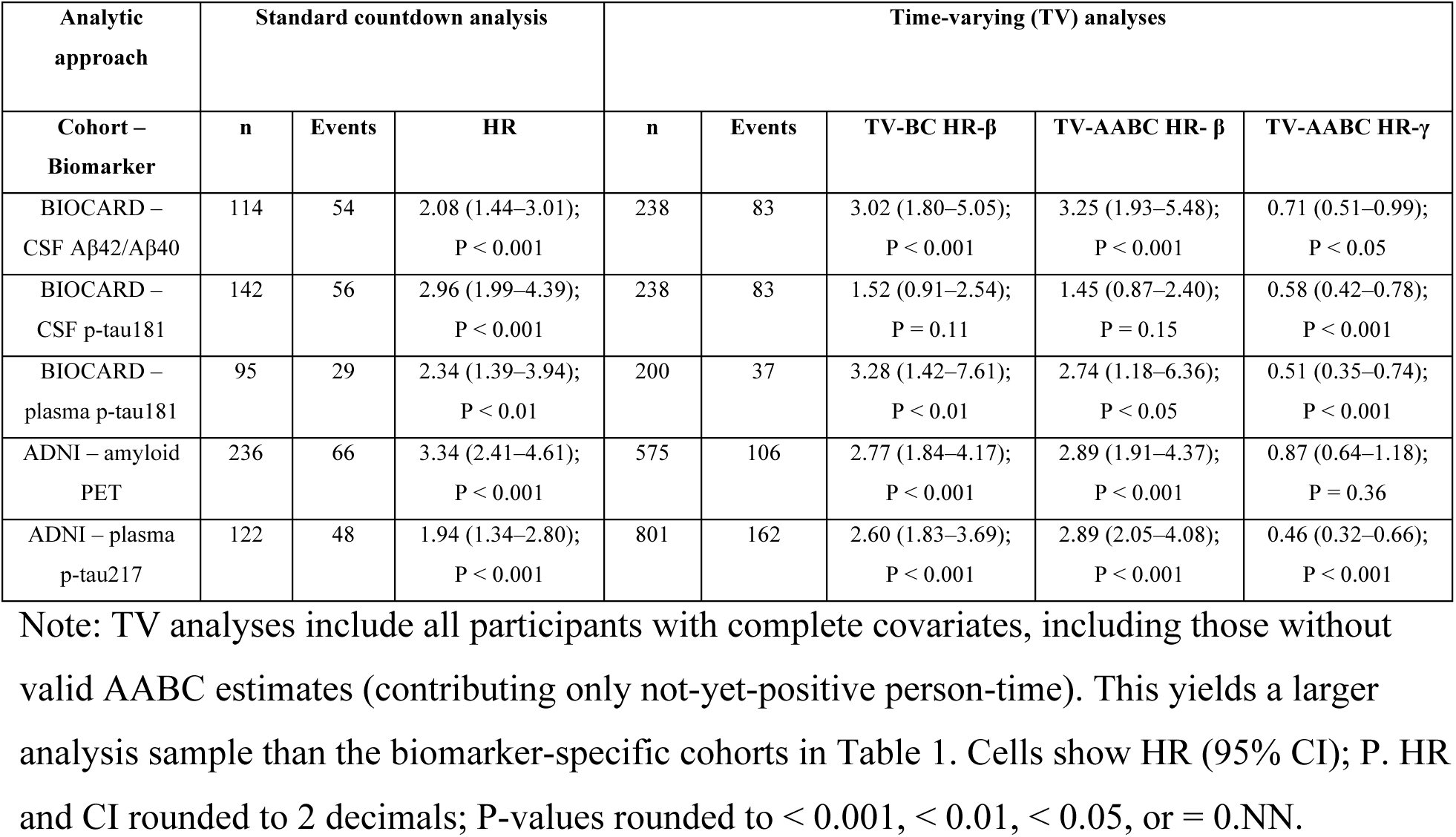
Main results: standard countdown vs. TV analyses.

| Analytic approach | Standard countdown analysis |  |  | Time-varying (TV) analyses |  |  |  |  |
| --- | --- | --- | --- | --- | --- | --- | --- | --- |
| Cohort – Biomarker | n | Events | HR | n | Events | TV-BC HR-β | TV-AABC HR- β | TV-AABC HR-γ |
| BIOCARD – CSF Aβ42/Aβ40 | 114 | 54 | 2.08 (1.44–3.01);<br>P < 0.001 | 238 | 83 | 3.02 (1.80–5.05);<br>P < 0.001 | 3.25 (1.93–5.48);<br>P < 0.001 | 0.71 (0.51–0.99);<br>P < 0.05 |
| BIOCARD – CSF p-tau181 | 142 | 56 | 2.96 (1.99–4.39);<br>P < 0.001 | 238 | 83 | 1.52 (0.91–2.54);<br>P = 0.11 | 1.45 (0.87–2.40);<br>P = 0.15 | 0.58 (0.42–0.78);<br>P < 0.001 |
| BIOCARD – plasma p-tau181 | 95 | 29 | 2.34 (1.39–3.94);<br>P < 0.01 | 200 | 37 | 3.28 (1.42–7.61);<br>P < 0.01 | 2.74 (1.18–6.36);<br>P < 0.05 | 0.51 (0.35–0.74);<br>P < 0.001 |
| ADNI – amyloid PET | 236 | 66 | 3.34 (2.41–4.61);<br>P < 0.001 | 575 | 106 | 2.77 (1.84–4.17);<br>P < 0.001 | 2.89 (1.91–4.37);<br>P < 0.001 | 0.87 (0.64–1.18);<br>P = 0.36 |
| ADNI – plasma p-tau217 | 122 | 48 | 1.94 (1.34–2.80);<br>P < 0.001 | 801 | 162 | 2.60 (1.83–3.69);<br>P < 0.001 | 2.89 (2.05–4.08);<br>P < 0.001 | 0.46 (0.32–0.66);<br>P < 0.001 |

Note: TV analyses include all participants with complete covariates, including those without valid AABC estimates (contributing only not-yet-positive person-time). This yields a larger analysis sample than the biomarker-specific cohorts in Table 1. Cells show HR (95% CI); P. HR and CI rounded to 2 decimals; P-values rounded to < 0.001, < 0.01, < 0.05, or = 0.NN.

### 3.5 Concerns with the standard biomarker-clock analysis approach

The most concerning property of the countdown paradox is its impact on inflated false positive rates and erroneously estimated effect directionality. This means published findings of significant associations between AABC and remaining time may reflect a structural artifact rather than a biological relationship. In practice, researchers cannot know a priori the presence or absence, or the direction of true effects, which limits the reliability of the standard countdown analysis for drawing clinically relevant insights surrounding biomarker-clock events or AABC.

## 4. Discussion

We compared, both theoretically and empirically, the standard countdown analytic approach used in many biomarker-clock studies with a time-varying effect methodology. Our results indicate the standard countdown analysis of age at a biomarker-clock event, which regresses remaining time to symptom onset on AABC, can produce structurally artifactual results that may be misleading. This countdown paradox arises from the mechanical relationship between predictor and outcome, rather than from estimation error or confounding; it is also distinct from the collider (selection) bias that can affect such analyses. More specifically, the remaining-time outcome introduces a structural negative covariance with the predictor, even when AABC and age at symptom onset are independent. The time-varying effect analysis avoids this structural coupling by operating on the natural age time scale and leveraging all person-time for estimation. Our simulations illustrate this paradox, and analyses of five biomarkers in the BIOCARD and ADNI cohorts are consistent with it.

Our findings have implications for analyses that have used the standard countdown analytic approach^4–6^. The finding that later biomarker positivity is associated with shorter remaining time to the outcome of interest may reflect the countdown-paradox artifact rather than a biologically meaningful association. The concern is that such a finding could invert the prognostic message—implying that later biomarker positivity portends faster progression, whereas the time-varying analytic approach may instead indicate that earlier positivity carries the greater risk.

It is important to emphasize that this does not mean the underlying biomarker science is invalid—amyloid positivity is a risk factor for clinical progression, as supported by multiple analyses, including the time-varying approach applied here to the BIOCARD and ADNI cohorts. Rather, we recommend that analyses examining how age at positivity influences the risk of progression to MCI or age at MCI onset require re-evaluation with methods that avoid this structural artifact. Importantly, covariate effects from published countdown analyses (e.g., *APOE* ε4, sex, education) remain valid. The structural bias is specific to the age-at-positivity predictor because it is the variable entangled with the time origin.

Recent independent work has sparked discussions^29, 30^ about structural artifacts in biomarker-clock analyses. As Insel and Donohue note^29^, regressing age at symptom onset directly on the AABC among individuals with an estimated AABC before symptom onset likewise induces an inaccurate positive association. This is the collider (selection) bias widely recognized in epidemiology^31, 32^ and is a separate issue from the structural covariance problem that drives the countdown paradox. The collider bias affects both the analysis noted by Insel and Donohue, which regresses age at symptom onset on AABC, and the standard countdown analysis discussed in this work, because both analyses are restricted to participants with AABC before symptom onset. An illustration of this related but distinct underlying structure is given in Supplementary Note 1. While our work focused on the underlying mechanism of the countdown paradox, we also provide a general methodological approach for analyzing the effect of biomarker-clock events and AABCs on disease outcomes.

These findings suggest three practical recommendations in terms of next steps. First, try using the time-varying effect analysis when the research question involves the age at a biomarker-clock event as a predictor and a time to disease outcome (e.g., age at MCI symptom onset) as the outcome of interest. Second, report both the TV-BC formulation (does experiencing the biomarker-clock event increase the hazard?) and the TV-AABC formulation (does AABC additionally modulate the hazard among positive individuals?). Third, interpret TV results on the age scale while controlling for the effects of other covariates. The HR-β compares the hazard of the outcome of interest on the age scale between positive and not-yet-positive individuals; the HR-γ compares, among positive individuals, the hazard for those whose biomarker-clock event occurred at a one-standard-deviation earlier versus later age, isolating the prognostic value of AABC itself.

Several limitations of our study should be noted. First, biomarker-clock status and AABC reflect each subject’s own evolving biomarker trajectory rather than an externally assigned exposure, which complicates causal interpretation. Second, although the structural negative covariance between age at positivity and remaining time is theoretically founded, the simulation study used relatively simple models. Third, the cross-cohort interpretation of TV-AABC results is limited by non-overlapping AABC distributions; HR-β at any single centering point is not directly comparable across cohorts. Fourth, when most participants are already biomarker positive at the analytic baseline, when there is limited variability in AABC, or when there is a small effect size in relation to the outcome of interest, the time-varying analysis may have limited power. Fifth, both the BIOCARD and ADNI cohorts are predominantly non-Hispanic White and highly educated, which limits generalizability. Although the countdown paradox is a structural property of the standard countdown analysis and therefore applies across cohorts, the results we report require replication in larger, more diverse cohorts. Finally, the TV-AABC analyses treat the SILA-estimated ages at biomarker positivity as given and do not propagate the uncertainty of the SILA trajectory fits into the Cox model; parametric bootstrap or measurement-error models could address this in future work.

In summary, the standard biomarker-clock analytic approach can generate inaccurate associations in remaining-time analyses and may reverse the apparent direction of true effects, which in turn inverts the prognostic message that biomarker-clock studies would convey to patients and clinicians. The time-varying effect analysis avoids this coupling by operating on the age time scale and using all person-time. Prior findings using the standard analytic approach may benefit from re-evaluation with the time-varying analysis we describe.

## Data availability

This study is a secondary analysis of existing cohort data available under controlled access. BIOCARD data are available to qualified investigators upon application to the BIOCARD study. ADNI data are available from the ADNI database (adni.loni.usc.edu) upon registration and approval. No new data were generated in this study. All simulation code and analysis scripts are available at https://github.com/dzhuyx/CountdownParadox; the simulation-study results are reproducible from the repository.

## Acknowledgements

We thank the participants and staff of the BIOCARD and ADNI studies. Part of the data used in preparation of this article were obtained from the Alzheimer’s Disease Neuroimaging Initiative (ADNI) database (adni.loni.usc.edu). As such, the investigators within the ADNI contributed to the design and implementation of ADNI and/or provided data but did not participate in analysis or writing of this report. A complete listing of ADNI investigators can be found at: http://adni.loni.usc.edu/wp-content/uploads/how_to_apply/ADNI_Acknowledgement_List.pdf

We thank Megan Clark for writing assistance in revising the manuscript. The authors used large language model tools (Claude and Claude Code; Anthropic) to assist with copy-editing, reference formatting, and internal-consistency checking of the manuscript, and with writing and revising analysis and figure-generation code; all such output was reviewed and edited by the authors, who take full responsibility for the content.

## Funding

This work was supported by the National Institute on Aging of the National Institutes of Health. Grant U19AG033655, which supports the BIOCARD study, provided support to Y.Z., C.P., A.S., A.M., M.A., and MC.W.; grant R01AG088637 provided support to Y.Z. and MC.W. The content is solely the responsibility of the authors and does not necessarily represent the official views of the National Institutes of Health.

## Competing interests

The authors report no competing interests.

## Supplementary material

Supplementary material is available online with this preprint.

## Notes

### Competing Interest Statement

The authors have declared no competing interest.

### Author Declarations

This study is a secondary analysis of de-identified data from two existing studies of human participants and did not involve the collection of any new data. The BIOCARD study was approved by the Institutional Review Board at Johns Hopkins, and all BIOCARD participants provided written informed consent. The Alzheimer's Disease Neuroimaging Initiative (ADNI) was approved by the institutional review boards of all participating institutions, and written informed consent was obtained from all ADNI participants or their authorized representatives.

### Summary of Updates

Title shortened; abstract expanded from about 148 to about 387 words to report the main quantitative results (standard-analysis false-positive rates up to 100 percent; empirical hazard ratios 1.94 to 3.34, all with P values below 0.01; reversal of the apparent age effect under the time-varying analysis), and keywords added; Methods reorganized, with the standard, time-varying, and simulation descriptions grouped under a Statistical analysis section and an Ethics statement added; a redundant Results overview subsection removed and the cross-cohort comparison revised for accuracy; text copy-edited and trimmed and coauthor edits incorporated; Data availability, Funding, and Competing interests sections added, and Acknowledgements expanded to disclose writing assistance and use of large language model tools, all reviewed by the authors; references reformatted to numbered AMA style (32 references) with author lists and publication years verified against source records; Figure 2 revised (legend labels updated to HR-beta and HR-gamma for consistency with the text); Figure 3 revised (onset-risk curves shown at the model-centering mean age at biomarker-clock event, with a note reconciling this value with the descriptive mean in the tables); alternative text added to all figure legends; Tables updated to match the revised content, with a reconciling footnote added to the degeneracy-assessment table; supplemental files updated (supplementary figures relabeled with the same hazard-ratio labels). No analyses, results, or conclusions were changed; the author list, license (CC BY-NC-ND), and subject area (Neurology) are unchanged.

