## Supplemental notes, tables, and figures for "The countdown paradox: time-varying analysis of biomarker-clock age and symptom onset"

### Supplementary contents

**Supplementary Note 1.** Mathematical derivation of the countdown paradox

**Supplementary Note 2.** Simulation study design

**Supplementary Note 3.** Study 1 simulation full results

**Supplementary Note 4.** Study 2 simulation full results

**Supplementary Table 1.** Study 1 complete results

**Supplementary Table 2.** Study 2 effect size results

**Supplementary Table 3.** Study 2 sample size results

**Supplementary Table 4.** Study 2 trajectory heterogeneity results

**Supplementary Table 5.** Study 2 disease progression rate results

**Supplementary Table 6.** Study 2 measurement precision results

**Supplementary Table 7.** Study 2 cross-product scenario results

**Supplementary Table 8.** Full model coefficients for all Cox PH models

**Supplementary Figure 1.** Type I error across Study 1 scenarios

**Supplementary Figure 2.** HR distributions under the null

### Supplementary Note 1. Mathematical derivation of the countdown paradox

#### Setup and notation

Let T denote each participant's age at the clinical event (onset of mild cognitive impairment, MCI), and let Y(t) denote the longitudinal biomarker trajectory at age t. We consider the null scenario that the biomarker trajectory carries no information about disease timing: the entire trajectory $\{Y(t): t \geq 0\}$ is independent of T, which we write as $Y(\cdot) \perp T$. The age at biomarker positivity (hereafter age-at-biomarker-clock event, AABC, when used as a covariate), Z, is obtained by applying a functional $g(\cdot)$ to the trajectory: $Z = g(Y(\cdot))$; examples include the simple case of a subject's age when the biomarker first reaches a cut-off threshold, the age estimated by applying the Sampled Iterative Local Approximation (SILA) algorithm, and the age estimated by inverting a fitted linear mixed-model trajectory at a pre-specified threshold as adopted in our simulation studies. Because Z is a function that uses only biomarker data $Y(\cdot)$ and never T, the independence relationship $Y(\cdot) \perp T$ transfers directly to $Z \perp T$; this implication holds regardless of which functional g is chosen. Define $R = T - Z$, the remaining time from biomarker positivity to clinical onset.

#### The structural negative covariance

Even under the strong null $Y(\cdot) \perp T$, which, as shown above, implies $Z \perp T$, the remaining time R is negatively correlated with Z by construction:

$$\mathrm{Cov}(Z, R) = \mathrm{Cov}(Z, T - Z) = \mathrm{Cov}(Z, T) - \mathrm{Var}(Z) = -\mathrm{Var}(Z) < 0.$$

A Cox proportional hazards model fit to (R, Z) inherits this structural negative association: larger Z (later biomarker positivity) is paired by construction with smaller R (less remaining time, equivalently higher hazard on the remaining-time scale), so the estimated hazard ratio for Z tends to exceed 1. Given sufficient sample size, the standard test of $H_0$: HR = 1 rejects at any pre-specified significance level, even though the biomarker trajectory is by assumption independent of disease onset. This is the countdown paradox: a guaranteed component of spurious association between age at biomarker positivity and remaining time, statistically significant at large N, driven entirely by the structural identity $\mathrm{Cov}(Z, T - Z) = -\mathrm{Var}(Z)$, rather than by any biological link between biomarker and disease.

**Selection-induced positive association when regressing onset age on AABC**

A related artifact, noted by Insel and Donohue,[29] arises if the age at clinical onset T is regressed directly on the AABC, Z, among participants for whom an AABC is estimated to precede symptom onset, i.e. conditional on the selection event $\{T > Z\}$. Even though $Z \perp T$ unconditionally ($\mathrm{Cov}(Z, T) = 0$), $\{T > Z\}$ is a function of both variables, so conditioning on it is a collider (selection) operation that induces dependence; $\mathrm{Corr}(T, Z \mid T > Z)$ is generally nonzero and depends on the marginals, but is nonnegative in this specific instance, as we illustrate below.

By the law of total covariance, conditioning on T within the selected set,

$$\mathrm{Cov}(T, Z \mid T > Z) = \mathrm{Cov}(T, E[Z \mid T, T > Z] \mid T > Z) + E[\mathrm{Cov}(T, Z \mid T, T > Z) \mid T > Z].$$

The second term is zero because T is fixed in the inner conditioning. Given $T = t$, the event $\{T > Z\}$ reduces to $\{Z < t\}$, so $E[Z \mid T = t, T > Z] = E[Z \mid Z < t]$, the upper-truncated mean of Z, which is nondecreasing in t. The covariance of T with a nondecreasing function of T is nonnegative, hence

$$\mathrm{Corr}(T, Z \mid T > Z) \geq 0,$$

with equality only in degenerate cases. Regressing onset age directly on AABC among biomarker-positive subjects therefore yields a spurious positive association.

Note that the collider bias illustrated above is also present in the standard countdown analysis, layered on top of the induced negative correlation between remaining time and AABC, because the standard countdown analysis is also restricted to participants with AABC estimated before symptom onset.

### TV-BC hazard model

The TV-BC (time-varying biomarker-clock) formulation specifies the age-specific hazard of T as:

$$\lambda(t \mid Z, X) = \lambda_0(t) \exp\{\beta \cdot 1[t \geq Z] + X^T\alpha\},$$

where $1[t \geq Z]$ is the time-varying indicator that age t has crossed the biomarker-positivity threshold age Z, X is a vector of baseline covariates, and $\lambda_0(t)$ is the (unspecified) baseline hazard. The coefficient β corresponds to the log hazard ratio comparing positive vs. not-yet-positive person-time at any given age. The TV-BC model is the natural generalization of a baseline-positivity Cox model and asks whether biomarker positivity increases the hazard of MCI onset.

### TV-AABC interaction hazard model

The TV-AABC (time-varying age-at-biomarker-clock) interaction model extends TV-BC by allowing the hazard of T among positive person-time to vary with age at positivity:

$$\lambda(t \mid Z, X) = \lambda_0(t) \exp\{\beta \cdot 1[t \geq Z] + \gamma \cdot A \cdot 1[t \geq Z] + X^T\alpha\},$$

where A is the z-scored age at biomarker positivity (defined only when $1[t \geq Z] = 1$). The term $\gamma \cdot A \cdot 1[t \geq Z]$ is an interaction between positivity status and AABC: it permits the hazard ratio for positivity to differ across the AABC distribution among positive subjects. HR-β = exp(β) is the hazard ratio at the mean AABC (A = 0), and HR-γ = exp(γ) is the multiplicative change in hazard per 1 SD increase in AABC among positive person-time. Under $Z \perp T$, both β and γ are 0 in expectation.

### Supplementary Note 2. Simulation study design

#### Study 1: Direct generation

Study 1 demonstrates the countdown paradox in its simplest form, using simple distributions to generate AABC and age at MCI onset directly, without biomarker trajectory modeling. This isolates the structural mechanism from any complexity of the estimation pipeline.

Fifteen scenarios were tested. Nine scenarios (S1–S9) used varied distributional families for AABC and age at MCI onset: Uniform–Uniform (S1), Normal–Normal (S2), Normal–Weibull (S3), Gamma–Weibull (S4), Beta–Exponential (S5), plus variants testing light censoring (S6, entry 50–60, 35-year follow-up), heavy censoring (S7, entry 60–75, 12-year follow-up), and high AABC variance (S8, AABC drawn from a normal distribution with mean 62 and SD 10). All used n = 500. One BIOCARD-calibrated scenario (S9) used AABC drawn from a normal distribution with mean 53 and SD 10, onset age from a normal distribution with mean 73 and SD 10, entry ages 40–65, and 20-year follow-up. Six additional sample-size variants tested S1 and S9 (BIOCARD-calibrated scenario) at n = 150, 200, and 1,000. For each scenario, 1,000 replications were generated using an oversample–filter–subsample design to ensure the target sample size after applying study entry and follow-up criteria.

**Study 2: LMM-based trajectory generation**

Study 2 uses a realistic data-generating process that mirrors the real data analysis pipeline. Biomarker trajectories are simulated from a linear mixed model; age at positivity is estimated by inverting and extrapolating the subject-specific trajectory, which is constructed with the population fixed-effect intercept and slope plus the subject's empirical-Bayes random-effect BLUPs, to the positivity threshold; and MCI onset is generated conditional on the baseline biomarker value.

Each subject's biomarker trajectory was generated from a linear function of age with subject-specific intercepts and slopes drawn from a bivariate normal distribution plus noise. The base scenario used the following default parameter values (varied across scenarios as described below): random-intercept standard deviation 0.5, random-slope standard deviation 0.008, intercept–slope correlation −0.3, measurement error standard deviation 0.25, fixed intercept −1.5, fixed slope 0.05, and positivity threshold 1.5. Age at biomarker positivity was estimated by fitting a linear mixed model to simulated longitudinal data and inverting the subject-specific trajectory at the positivity threshold. Clinical onset was generated by adding an exponentially distributed waiting time to the study entry age, with the hazard rate depending on the subject's baseline biomarker level through a proportional hazards specification, where the log-hazard increased linearly with the centered baseline biomarker level scaled by a true effect parameter that controlled the association between the biomarker and disease risk. Two base study-design scenarios were used: a pedagogical design (PED; entry ages 50–65, biennial visits, baseline hazard 0.03 per year, n = 500) and a BIOCARD-resembling design (BIO; entry ages 40–70, quadrennial visits, baseline hazard 0.02 per year, n = 500).

Each of the 59 scenarios (19 null and 40 non-null) started from one of the two base scenarios and varied one or more parameters. The scenarios were organized into six categories. (1) Effect size variation (14 scenarios; 12 non-null + 2 null at β_true = 0): the true effect size ranged from −1.0 to +1.0 in both PED and BIO families, testing null, mild (±0.3), moderate (±0.5), and strong (±1.0) effects. (2) Sample size variation (10 scenarios; 5 non-null + 5 paired null twins): sample sizes from 150 to 2,000 with a true effect size of 0.5, each paired with a null twin at β_true = 0. (3) Trajectory heterogeneity (8 scenarios; 4 non-null + 4 null): the random-slope standard deviation ranged from 0.004 (low) to 0.016 (high), altering the variance of age at positivity. (4) Disease progression rate (8 scenarios; 4 non-null + 4 null): the baseline hazard ranged from 0.015 to 0.05 per year. (5) Measurement precision (8 scenarios; 4 non-null + 4 null): visit intervals from 1 to 4 years and measurement error standard deviation from 0.25 to 0.5. (6) Cross-product combinations (11 non-null scenarios): two or more parameters varied simultaneously—for example, strong effects with large samples, high heterogeneity, or fast disease progression—including a maximum-power scenario (true effect size = 1.0, n = 2,000, baseline hazard = 0.05).

#### Evaluation metrics

For each scenario and analytic approach, we computed the following metrics across 1,000 replications: (1) rejection rate—representing type I error under the null and statistical power under the alternative; (2) mean and median hazard ratio; (3) mean log hazard ratio as a measure of bias; (4) direction accuracy: the proportion of replications in which the estimated hazard ratio was in the correct direction (applicable only under the alternative).

### Supplementary Note 3. Study 1 full results (direct generation)

**Supplementary Table 1.** Study 1 complete results (15 scenarios).

| Scenario | N | Analytic approach | AABC variance | Event rate (%) | Type I error | Mean HR | SD HR |
|---|---|---|---|---|---|---|---|
| S1: Uniform-Uniform | 500 | Standard countdown | 52.1 | 71.9% | 1.000 | 1.12 | 0.011 |
| S1: Uniform-Uniform | 500 | TV-BC | 52.1 | 71.9% | 0.058 | 1.02 | 0.175 |
| S1: Uniform-Uniform | 500 | TV-AABC β | 52.1 | 71.9% | 0.055 | 1.02 | 0.177 |
| S1: Uniform-Uniform | 500 | TV-AABC γ | 52.1 | 71.9% | 0.049 | 1.00 | 0.062 |
| S9: BIOCARD-calibrated | 500 | Standard countdown | 96.0 | 46.2% | 1.000 | 1.14 | 0.012 |
| S9: BIOCARD-calibrated | 500 | TV-BC | 96.0 | 46.2% | 0.051 | 1.04 | 0.240 |
| S9: BIOCARD-calibrated | 500 | TV-AABC β | 96.0 | 46.2% | 0.047 | 1.04 | 0.241 |
| S9: BIOCARD-calibrated | 500 | TV-AABC γ | 96.0 | 46.2% | 0.045 | 1.00 | 0.071 |
| S9, n=1000: BIOCARD-calibrated (n=1000) | 1000 | Standard countdown | 96.1 | 46.4% | 1.000 | 1.14 | 0.009 |
| S9, n=1000: BIOCARD-calibrated (n=1000) | 1000 | TV-BC | 96.1 | 46.4% | 0.049 | 1.02 | 0.166 |
| S9, n=1000: BIOCARD-calibrated (n=1000) | 1000 | TV-AABC β | 96.1 | 46.4% | 0.049 | 1.02 | 0.166 |
| S9, n=1000: BIOCARD-calibrated (n=1000) | 1000 | TV-AABC γ | 96.1 | 46.4% | 0.051 | 1.00 | 0.050 |
| S9, n=150: BIOCARD-calibrated (n=150) | 150 | Standard countdown | 95.9 | 46.4% | 1.000 | 1.14 | 0.022 |
| S9, n=150: BIOCARD-calibrated (n=150) | 150 | TV-BC | 95.9 | 46.4% | 0.044 | 1.14 | 0.504 |
| S9, n=150: BIOCARD- | 150 | TV-AABC | 95.9 | 46.4% | 0.042 | 1.13 | 0.504 |

| | | | | | | | |
|---|---|---|---|---|---|---|---|
| calibrated (n=150) | | β | | | | | |
| S9, n=150: BIOCARD-calibrated (n=150) | 150 | TV-AABC γ | 95.9 | 46.4% | 0.051 | 1.01 | 0.133 |
| S9, n=200: BIOCARD-calibrated (n=200) | 200 | Standard countdown | 96.1 | 46.2% | 1.000 | 1.14 | 0.020 |
| S9, n=200: BIOCARD-calibrated (n=200) | 200 | TV-BC | 96.1 | 46.2% | 0.047 | 1.10 | 0.411 |
| S9, n=200: BIOCARD-calibrated (n=200) | 200 | TV-AABC β | 96.1 | 46.2% | 0.044 | 1.09 | 0.412 |
| S9, n=200: BIOCARD-calibrated (n=200) | 200 | TV-AABC γ | 96.1 | 46.2% | 0.048 | 1.01 | 0.115 |
| S1, n=1000: Uniform-Uniform (n=1000) | 1000 | Standard countdown | 52.2 | 71.9% | 1.000 | 1.12 | 0.008 |
| S1, n=1000: Uniform-Uniform (n=1000) | 1000 | TV-BC | 52.2 | 71.9% | 0.046 | 1.00 | 0.118 |
| S1, n=1000: Uniform-Uniform (n=1000) | 1000 | TV-AABC β | 52.2 | 71.9% | 0.049 | 1.00 | 0.119 |
| S1, n=1000: Uniform-Uniform (n=1000) | 1000 | TV-AABC γ | 52.2 | 71.9% | 0.048 | 1.00 | 0.043 |
| S1, n=150: Uniform-Uniform (n=150) | 150 | Standard countdown | 52.2 | 71.8% | 1.000 | 1.12 | 0.020 |
| S1, n=150: Uniform-Uniform (n=150) | 150 | TV-BC | 52.2 | 71.8% | 0.042 | 1.09 | 0.341 |
| S1, n=150: Uniform-Uniform (n=150) | 150 | TV-AABC β | 52.2 | 71.8% | 0.038 | 1.08 | 0.343 |
| S1, n=150: Uniform-Uniform (n=150) | 150 | TV-AABC γ | 52.2 | 71.8% | 0.048 | 1.00 | 0.115 |
| S1, n=200: Uniform-Uniform (n=200) | 200 | Standard countdown | 51.8 | 71.7% | 1.000 | 1.12 | 0.018 |
| S1, n=200: Uniform- | 200 | TV-BC | 51.8 | 71.7% | 0.056 | 1.05 | 0.304 |

| | | | | | | | |
|---|---|---|---|---|---|---|---|
| Uniform (n=200) | | | | | | | |
| S1, n=200: Uniform-Uniform (n=200) | 200 | TV-AABC β | 51.8 | 71.7% | 0.056 | 1.05 | 0.307 |
| S1, n=200: Uniform-Uniform (n=200) | 200 | TV-AABC γ | 51.8 | 71.7% | 0.047 | 1.00 | 0.099 |
| S2: Normal-Normal | 500 | Standard countdown | 24.8 | 80.5% | 1.000 | 1.17 | 0.016 |
| S2: Normal-Normal | 500 | TV-BC | 24.8 | 80.5% | 0.048 | 1.05 | 0.282 |
| S2: Normal-Normal | 500 | TV-AABC β | 24.8 | 80.5% | 0.047 | 1.05 | 0.282 |
| S2: Normal-Normal | 500 | TV-AABC γ | 24.8 | 80.5% | 0.055 | 1.00 | 0.053 |
| S3: Normal-Weibull | 500 | Standard countdown | 24.9 | 61.7% | 1.000 | 1.09 | 0.015 |
| S3: Normal-Weibull | 500 | TV-BC | 24.9 | 61.7% | 0.046 | 1.02 | 0.241 |
| S3: Normal-Weibull | 500 | TV-AABC β | 24.9 | 61.7% | 0.046 | 1.02 | 0.240 |
| S3: Normal-Weibull | 500 | TV-AABC γ | 24.9 | 61.7% | 0.045 | 1.00 | 0.063 |
| S4: Gamma-Weibull | 500 | Standard countdown | 24.3 | 87.4% | 1.000 | 1.16 | 0.016 |
| S4: Gamma-Weibull | 500 | TV-BC | 24.3 | 87.4% | 0.049 | 1.03 | 0.215 |
| S4: Gamma-Weibull | 500 | TV-AABC β | 24.3 | 87.4% | 0.050 | 1.03 | 0.215 |
| S4: Gamma-Weibull | 500 | TV-AABC γ | 24.3 | 87.4% | 0.047 | 1.00 | 0.051 |
| S5: Beta-Exponential | 500 | Standard countdown | 31.2 | 61.3% | 1.000 | 1.08 | 0.013 |
| S5: Beta-Exponential | 500 | TV-BC | 31.2 | 61.3% | 0.040 | 1.12 | 0.413 |
| S5: Beta-Exponential | 500 | TV-AABC | 31.2 | 61.3% | 0.039 | 1.12 | 0.413 |

| | | | | | | | |
|---|---|---|---|---|---|---|---|
| | | β | | | | | |
| S5: Beta-Exponential | 500 | TV-AABC γ | 31.2 | 61.3% | 0.050 | 1.00 | 0.060 |
| S6: Light Censoring | 500 | Standard countdown | 24.9 | 97.8% | 1.000 | 1.14 | 0.013 |
| S6: Light Censoring | 500 | TV-BC | 24.9 | 97.8% | 0.049 | 1.04 | 0.244 |
| S6: Light Censoring | 500 | TV-AABC β | 24.9 | 97.8% | 0.053 | 1.04 | 0.245 |
| S6: Light Censoring | 500 | TV-AABC γ | 24.9 | 97.8% | 0.055 | 1.00 | 0.048 |
| S7: Heavy Censoring | 500 | Standard countdown | 24.8 | 64.1% | 1.000 | 1.21 | 0.019 |
| S7: Heavy Censoring | 500 | TV-BC | 24.8 | 64.1% | 0.047 | 1.11 | 0.382 |
| S7: Heavy Censoring | 500 | TV-AABC β | 24.8 | 64.1% | 0.046 | 1.11 | 0.383 |
| S7: Heavy Censoring | 500 | TV-AABC γ | 24.8 | 64.1% | 0.044 | 1.00 | 0.058 |
| S8: High AABC variance | 500 | Standard countdown | 95.1 | 80.6% | 1.000 | 1.17 | 0.012 |
| S8: High AABC variance | 500 | TV-BC | 95.1 | 80.6% | 0.060 | 1.01 | 0.155 |
| S8: High AABC variance | 500 | TV-AABC β | 95.1 | 80.6% | 0.054 | 1.01 | 0.155 |
| S8: High AABC variance | 500 | TV-AABC γ | 95.1 | 80.6% | 0.055 | 1.00 | 0.056 |

Study 1 results confirm that the standard countdown analysis produces type I error inflation across all 15 scenarios tested (range 100%–100%), while TV-BC maintains type I error near 5%

(range 4.0–6.0%), TV-AABC $\beta$ also maintains valid type I error (range 3.8–5.6%), and TV-AABC $\gamma$ maintains nominal levels (range 4.4–5.5%).

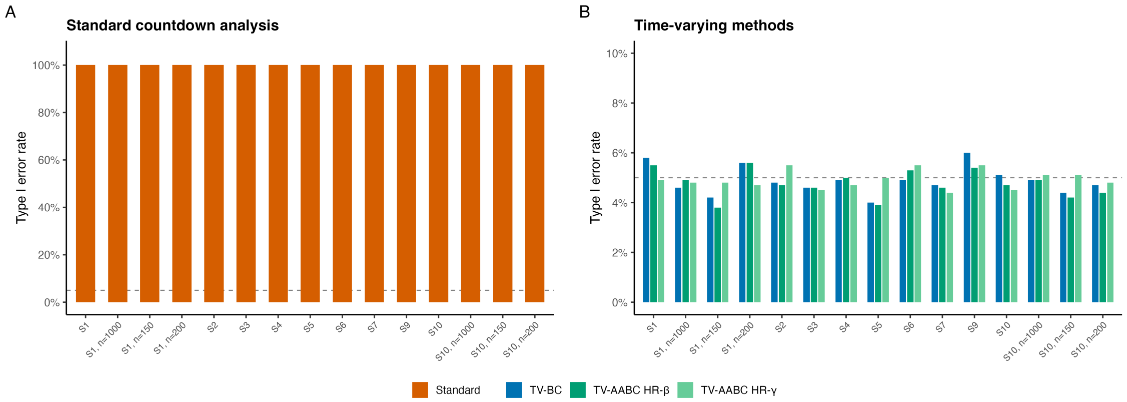


**Supplementary Figure 1.**

Type I error rates across all 15 Study 1 scenarios. Bar chart showing the rejection rate under the null (proportion of 1,000 replications with $P < 0.05$) for four analytic approaches: standard countdown (orange), TV-BC (blue), TV-AABC β (bluish green), and TV-AABC γ (light green). The dashed horizontal line indicates the nominal 5% level. The standard countdown analysis produces 100% type I error in all scenarios; all three time-varying approaches remain near 5%.

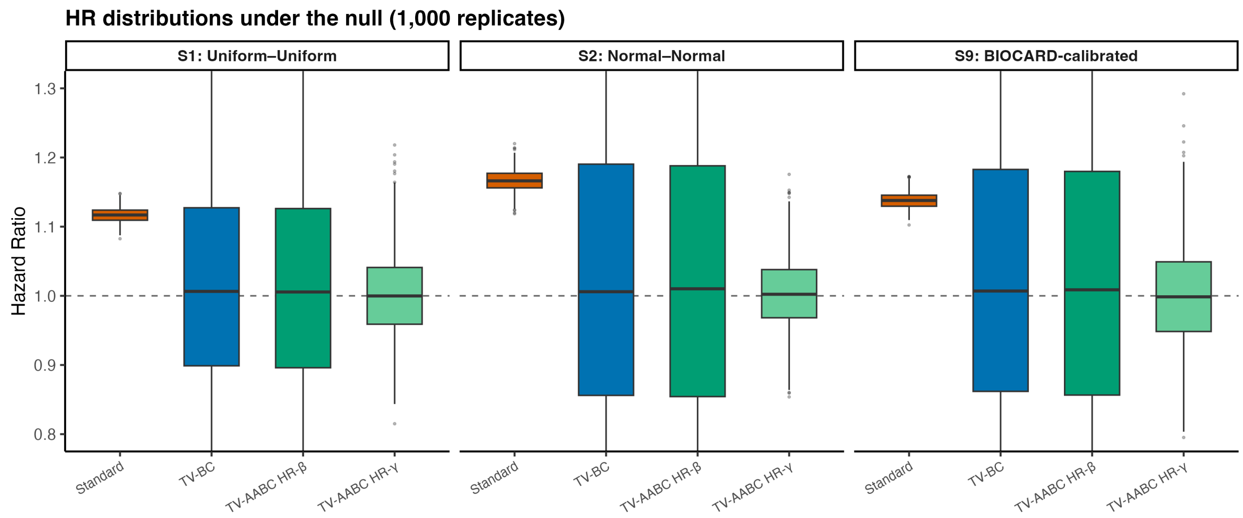


**Supplementary Figure 2.**

Hazard ratio distributions under the null for three representative Study 1 scenarios (S1: Uniform–Uniform; S2: Normal–Normal; S9: BIOCARD-calibrated), selected to span the range of distributional families tested. Boxplots of 1,000 hazard ratio replications are shown for each analytic approach: standard countdown (orange), TV-BC (blue), TV-AABC β (bluish green), and TV-AABC γ (light green). The standard countdown distribution is shifted well above 1.0; the time-varying approach distributions are centered at 1.0. The horizontal dashed line indicates HR = 1.0 (no association). The y-axis is restricted to 0.8–1.3 for visual clarity; 27% of TV-BC and TV-AABC β replications fall outside this range and are not displayed; all standard countdown and TV-AABC γ replications fall within the displayed range.

### Supplementary Note 4. Study 2 full results

Throughout this Note, scenarios are labelled by their base study-design scenario: PED denotes the pedagogical design (entry ages 50–65, biennial visits, baseline hazard 0.03 per year) and BIO denotes the BIOCARD-resembling design (entry ages 40–70, quadrennial visits, baseline hazard 0.02 per year). Full parameter specifications for each base scenario are given in Supplementary Note 2.

**Supplementary Table 2.** Study 2 effect size results. Family column: PED = pedagogical scenario; BIO = BIOCARD-resembling scenario.

| Scenario | Family | True effect size (β) | Analytic approach | Rejection rate | Mean HR | SD HR | % Correct direction |
|---|---|---|---|---|---|---|---|
| S3e-bio: Strong protective | BIO | -1.0 | Standard countdown | 0.972 | 1.05 | 0.011 | 100.0% |
| S3e-bio: Strong protective | BIO | -1.0 | TV-BC | 0.996 | 0.45 | 0.081 | 100.0% |
| S3e-bio: Strong protective | BIO | -1.0 | TV-AABC β | 0.923 | 0.53 | 0.108 | 99.9% |
| S3e-bio: Strong protective | BIO | -1.0 | TV-AABC γ | 0.946 | 1.52 | 0.179 | 100.0% |
| S3d-bio: Moderate protective | BIO | -0.5 | Standard countdown | 0.925 | 1.04 | 0.011 | 100.0% |
| S3d-bio: | BIO | -0.5 | TV-BC | 0.710 | 0.65 | 0.111 | 99.5% |

| | | | | | | | |
|---|---|---|---|---|---|---|---|
| Moderate protective | | | | | | | |
| S3d-bio: Moderate protective | BIO | -0.5 | TV-AABC β | 0.457 | 0.72 | 0.132 | 96.8% |
| S3d-bio: Moderate protective | BIO | -0.5 | TV-AABC γ | 0.542 | 1.26 | 0.140 | 98.5% |
| S3g-bio: Mild protective | BIO | -0.3 | Standard countdown | 0.880 | 1.03 | 0.011 | 100.0% |
| S3g-bio: Mild protective | BIO | -0.3 | TV-BC | 0.325 | 0.78 | 0.141 | 93.4% |
| S3g-bio: Mild protective | BIO | -0.3 | TV-AABC β | 0.200 | 0.84 | 0.161 | 83.4% |
| S3g-bio: Mild protective | BIO | -0.3 | TV-AABC γ | 0.275 | 1.17 | 0.126 | 92.1% |
| S3a-bio: Null effect | BIO | 0.0 | Standard countdown | 0.749 | 1.03 | 0.011 | — |
| S3a-bio: Null effect | BIO | 0.0 | TV-BC | 0.049 | 1.01 | 0.180 | — |
| S3a-bio: Null effect | BIO | 0.0 | TV-AABC β | 0.051 | 1.01 | 0.189 | — |
| S3a-bio: Null | BIO | 0.0 | TV-AABC γ | 0.060 | 1.02 | 0.105 | — |

| | | | | | | | |
|---|---|---|---|---|---|---|---|
| effect | | | | | | | |
| S3f-bio: Mild harmful | BIO | 0.3 | Standard countdown | 0.557 | 1.02 | 0.010 | 1.3% |
| S3f-bio: Mild harmful | BIO | 0.3 | TV-BC | 0.304 | 1.34 | 0.241 | 93.2% |
| S3f-bio: Mild harmful | BIO | 0.3 | TV-AABC β | 0.148 | 1.24 | 0.228 | 86.5% |
| S3f-bio: Mild harmful | BIO | 0.3 | TV-AABC γ | 0.269 | 0.88 | 0.087 | 92.6% |
| S3b-bio: Moderate harmful | BIO | 0.5 | Standard countdown | 0.393 | 1.02 | 0.010 | 3.2% |
| S3b-bio: Moderate harmful | BIO | 0.5 | TV-BC | 0.720 | 1.62 | 0.290 | 99.7% |
| S3b-bio: Moderate harmful | BIO | 0.5 | TV-AABC β | 0.394 | 1.42 | 0.258 | 97.2% |
| S3b-bio: Moderate harmful | BIO | 0.5 | TV-AABC γ | 0.662 | 0.80 | 0.078 | 99.5% |
| S3c-bio: Strong harmful | BIO | 1.0 | Standard countdown | 0.079 | 1.01 | 0.008 | 25.5% |
| S3c-bio: Strong harmful | BIO | 1.0 | TV-BC | 0.999 | 2.51 | 0.487 | 100.0% |

| | | | | | | | |
|---|---|---|---|---|---|---|---|
| S3c-bio: Strong harmful | BIO | 1.0 | TV-AABC β | 0.896 | 1.87 | 0.365 | 100.0% |
| S3c-bio: Strong harmful | BIO | 1.0 | TV-AABC γ | 0.999 | 0.62 | 0.060 | 100.0% |
| S3e-ped: Strong protective | PED | -1.0 | Standard countdown | 1.000 | 1.08 | 0.012 | 100.0% |
| S3e-ped: Strong protective | PED | -1.0 | TV-BC | 1.000 | 0.44 | 0.063 | 100.0% |
| S3e-ped: Strong protective | PED | -1.0 | TV-AABC β | 0.999 | 0.49 | 0.075 | 100.0% |
| S3e-ped: Strong protective | PED | -1.0 | TV-AABC γ | 0.980 | 1.53 | 0.161 | 100.0% |
| S3d-ped: Moderate protective | PED | -0.5 | Standard countdown | 1.000 | 1.07 | 0.011 | 100.0% |
| S3d-ped: Moderate protective | PED | -0.5 | TV-BC | 0.845 | 0.65 | 0.098 | 99.8% |
| S3d-ped: Moderate protective | PED | -0.5 | TV-AABC β | 0.666 | 0.70 | 0.111 | 98.6% |
| S3d-ped: Moderate protective | PED | -0.5 | TV-AABC γ | 0.612 | 1.26 | 0.126 | 98.5% |
| S3g-ped: | PED | -0.3 | Standard | 1.000 | 1.06 | 0.011 | 100.0% |

| | | | | | | | |
|---|---|---|---|---|---|---|---|
| Mild protective | | | countdown | | | | |
| S3g-ped: Mild protective | PED | -0.3 | TV-BC | 0.446 | 0.77 | 0.114 | 96.9% |
| S3g-ped: Mild protective | PED | -0.3 | TV-AABC β | 0.292 | 0.81 | 0.126 | 93.0% |
| S3g-ped: Mild protective | PED | -0.3 | TV-AABC γ | 0.324 | 1.16 | 0.114 | 92.8% |
| S3a-ped: Null effect | PED | 0.0 | Standard countdown | 0.997 | 1.05 | 0.011 | — |
| S3a-ped: Null effect | PED | 0.0 | TV-BC | 0.057 | 1.02 | 0.161 | — |
| S3a-ped: Null effect | PED | 0.0 | TV-AABC β | 0.046 | 1.02 | 0.164 | — |
| S3a-ped: Null effect | PED | 0.0 | TV-AABC γ | 0.047 | 1.01 | 0.096 | — |
| S3f-ped: Mild harmful | PED | 0.3 | Standard countdown | 0.973 | 1.04 | 0.010 | 0.0% |
| S3f-ped: Mild harmful | PED | 0.3 | TV-BC | 0.425 | 1.35 | 0.210 | 97.1% |
| S3f-ped: Mild | PED | 0.3 | TV-AABC β | 0.272 | 1.27 | 0.206 | 91.0% |

| | | | | | | | |
|---|---|---|---|---|---|---|---|
| harmful | | | | | | | |
| S3f-ped: Mild harmful | PED | 0.3 | TV-AABC γ | 0.324 | 0.88 | 0.081 | 91.3% |
| S3b-ped: Moderate harmful | PED | 0.5 | Standard countdown | 0.928 | 1.04 | 0.010 | 0.0% |
| S3b-ped: Moderate harmful | PED | 0.5 | TV-BC | 0.852 | 1.63 | 0.266 | 99.7% |
| S3b-ped: Moderate harmful | PED | 0.5 | TV-AABC β | 0.608 | 1.47 | 0.245 | 98.7% |
| S3b-ped: Moderate harmful | PED | 0.5 | TV-AABC γ | 0.714 | 0.80 | 0.073 | 99.1% |
| S3c-ped: Strong harmful | PED | 1.0 | Standard countdown | 0.535 | 1.02 | 0.009 | 1.2% |
| S3c-ped: Strong harmful | PED | 1.0 | TV-BC | 1.000 | 2.53 | 0.448 | 100.0% |
| S3c-ped: Strong harmful | PED | 1.0 | TV-AABC β | 0.978 | 2.01 | 0.359 | 100.0% |
| S3c-ped: Strong harmful | PED | 1.0 | TV-AABC γ | 0.999 | 0.63 | 0.057 | 100.0% |

Supplementary Tables 3–7 present the full results for the remaining Study 2 sensitivity categories: sample size variation (Supplementary Table 3), trajectory heterogeneity (Supplementary Table 4), disease progression rate (Supplementary Table 5), measurement precision (Supplementary Table 6), and cross-product combinations (Supplementary Table 7). All tables follow the same format as Supplementary Table 2.

**Supplementary Table 3.** Study 2 sample size results. Family column: PED = pedagogical scenario; BIO = BIOCARD-resembling scenario.

| Scenario | Family | True effect size (β) | Analytic approach | Rejection rate | Mean HR | SD HR | % Correct direction |
|---|---|---|---|---|---|---|---|
| S3h-bio: BIOCARD n=150 | BIO | 0.5 | Standard countdown | 0.133 | 1.02 | 0.018 | 15.1% |
| S3h-bio: BIOCARD n=150 | BIO | 0.5 | TV-BC | 0.254 | 1.71 | 0.602 | 91.5% |
| S3h-bio: BIOCARD n=150 | BIO | 0.5 | TV-AABC β | 0.127 | 1.49 | 0.544 | 82.0% |
| S3h-bio: BIOCARD n=150 | BIO | 0.5 | TV-AABC γ | 0.258 | 0.81 | 0.146 | 90.1% |
| S3h-bio-null: BIOCARD n=150 (null) | BIO | 0.0 | Standard countdown | 0.306 | 1.03 | 0.019 | — |

| | | | | | | | |
|---|---|---|---|---|---|---|---|
| S3h-bio-null: BIOCARD n=150 (null) | BIO | 0.0 | TV-BC | 0.055 | 1.07 | 0.379 | — |
| S3h-bio-null: BIOCARD n=150 (null) | BIO | 0.0 | TV-AABC β | 0.059 | 1.09 | 0.406 | — |
| S3h-bio-null: BIOCARD n=150 (null) | BIO | 0.0 | TV-AABC γ | 0.058 | 1.04 | 0.201 | — |
| S3h-ped: Small sample | PED | 0.5 | Standard countdown | 0.549 | 1.04 | 0.016 | 1.1% |
| S3h-ped: Small sample | PED | 0.5 | TV-BC | 0.436 | 1.66 | 0.428 | 98.0% |
| S3h-ped: Small sample | PED | 0.5 | TV-AABC β | 0.271 | 1.50 | 0.392 | 92.6% |
| S3h-ped: Small sample | PED | 0.5 | TV-AABC γ | 0.334 | 0.81 | 0.117 | 94.0% |
| S3h-ped-null: Small sample (null) | PED | 0.0 | Standard countdown | 0.813 | 1.05 | 0.017 | — |

| | | | | | | | |
|---|---|---|---|---|---|---|---|
| S3h-ped-null: Small sample (null) | PED | 0.0 | TV-BC | 0.044 | 1.03 | 0.250 | — |
| S3h-ped-null: Small sample (null) | PED | 0.0 | TV-AABC β | 0.041 | 1.04 | 0.257 | — |
| S3h-ped-null: Small sample (null) | PED | 0.0 | TV-AABC γ | 0.046 | 1.02 | 0.155 | — |
| S3i-bio: BIOCARD large | BIO | 0.5 | Standard countdown | 0.648 | 1.02 | 0.007 | 0.7% |
| S3i-bio: BIOCARD large | BIO | 0.5 | TV-BC | 0.959 | 1.62 | 0.216 | 99.9% |
| S3i-bio: BIOCARD large | BIO | 0.5 | TV-AABC β | 0.721 | 1.42 | 0.189 | 99.8% |
| S3i-bio: BIOCARD large | BIO | 0.5 | TV-AABC γ | 0.930 | 0.79 | 0.055 | 100.0% |
| S3i-bio-null: BIOCARD large (null) | BIO | 0.0 | Standard countdown | 0.962 | 1.03 | 0.007 | — |
| S3i-bio-null: BIOCARD | BIO | 0.0 | TV-BC | 0.057 | 1.01 | 0.131 | — |

| | | | | | | | |
|---|---|---|---|---|---|---|---|
| large (null) | | | | | | | |
| S3i-bio-null: BIOCARD large (null) | BIO | 0.0 | TV-AABC β | 0.050 | 1.02 | 0.137 | — |
| S3i-bio-null: BIOCARD large (null) | BIO | 0.0 | TV-AABC γ | 0.060 | 1.01 | 0.074 | — |
| S3i-ped: Large sample | PED | 0.5 | Standard countdown | 1.000 | 1.04 | 0.007 | 0.0% |
| S3i-ped: Large sample | PED | 0.5 | TV-BC | 0.990 | 1.60 | 0.182 | 100.0% |
| S3i-ped: Large sample | PED | 0.5 | TV-AABC β | 0.856 | 1.44 | 0.169 | 100.0% |
| S3i-ped: Large sample | PED | 0.5 | TV-AABC γ | 0.934 | 0.80 | 0.052 | 100.0% |
| S3i-ped-null: Large sample (null) | PED | 0.0 | Standard countdown | 1.000 | 1.05 | 0.008 | — |
| S3i-ped-null: Large sample (null) | PED | 0.0 | TV-BC | 0.050 | 1.01 | 0.112 | — |
| S3i-ped-null: Large | PED | 0.0 | TV-AABC β | 0.056 | 1.02 | 0.116 | — |

| | | | | | | | |
|---|---|---|---|---|---|---|---|
| sample (null) | | | | | | | |
| S3i-ped-null: Large sample (null) | PED | 0.0 | TV-AABC γ | 0.050 | 1.01 | 0.068 | — |
| S3j-ped: Very large sample | PED | 0.5 | Standard countdown | 1.000 | 1.04 | 0.005 | 0.0% |
| S3j-ped: Very large sample | PED | 0.5 | TV-BC | 1.000 | 1.59 | 0.131 | 100.0% |
| S3j-ped: Very large sample | PED | 0.5 | TV-AABC β | 0.994 | 1.44 | 0.120 | 100.0% |
| S3j-ped: Very large sample | PED | 0.5 | TV-AABC γ | 0.999 | 0.80 | 0.036 | 100.0% |
| S3j-ped-null: Very large sample (null) | PED | 0.0 | Standard countdown | 1.000 | 1.05 | 0.005 | — |
| S3j-ped-null: Very large sample (null) | PED | 0.0 | TV-BC | 0.053 | 1.01 | 0.077 | — |
| S3j-ped-null: Very large | PED | 0.0 | TV-AABC β | 0.047 | 1.01 | 0.081 | — |

| | | | | | | | |
|---|---|---|---|---|---|---|---|
| sample (null) | | | | | | | |
| S3j-ped-null: Very large sample (null) | PED | 0.0 | TV-AABC γ | 0.063 | 1.01 | 0.048 | — |

**Supplementary Table 4.** Study 2 trajectory heterogeneity results. Family column: PED = pedagogical scenario; BIO = BIOCARD-resembling scenario.

| Scenario | Family | True effect size (β) | Analytic approach | Rejection rate | Mean HR | SD HR | % Correct direction |
|---|---|---|---|---|---|---|---|
| S3k-bio: Low heterogeneity | BIO | 0.5 | Standard countdown | 0.151 | 1.02 | 0.016 | 17.0% |
| S3k-bio: Low heterogeneity | BIO | 0.5 | TV-BC | 0.127 | 1.28 | 0.321 | 80.5% |
| S3k-bio: Low heterogeneity | BIO | 0.5 | TV-AABC β | 0.073 | 1.19 | 0.303 | 70.7% |
| S3k-bio: Low heterogeneity | BIO | 0.5 | TV-AABC γ | 0.199 | 0.91 | 0.081 | 87.9% |
| S3k-bio-null: Low | BIO | 0.0 | Standard countdown | 0.310 | 1.03 | 0.017 | — |

| | | | | | | | |
|---|---|---|---|---|---|---|---|
| heterogeneity (null) | | | | | | | |
| S3k-bio-null: Low heterogeneity (null) | BIO | 0.0 | TV-BC | 0.048 | 1.04 | 0.245 | — |
| S3k-bio-null: Low heterogeneity (null) | BIO | 0.0 | TV-AABC β | 0.047 | 1.05 | 0.260 | — |
| S3k-bio-null: Low heterogeneity (null) | BIO | 0.0 | TV-AABC γ | 0.051 | 1.03 | 0.096 | — |
| S3k-ped: Low heterogeneity | PED | 0.5 | Standard countdown | 0.299 | 1.02 | 0.015 | 6.1% |
| S3k-ped: Low heterogeneity | PED | 0.5 | TV-BC | 0.181 | 1.28 | 0.246 | 88.1% |
| S3k-ped: Low heterogeneity | PED | 0.5 | TV-AABC β | 0.105 | 1.21 | 0.240 | 79.0% |
| S3k-ped: Low heterogeneity | PED | 0.5 | TV-AABC γ | 0.202 | 0.92 | 0.078 | 84.4% |
| S3k-ped-null: Low heterogeneity (null) | PED | 0.0 | Standard countdown | 0.638 | 1.04 | 0.016 | — |
| S3k-ped- | PED | 0.0 | TV-BC | 0.047 | 1.03 | 0.199 | — |

| | | | | | | | |
|---|---|---|---|---|---|---|---|
| null: Low heterogeneity (null) | | | | | | | |
| S3k-ped-null: Low heterogeneity (null) | PED | 0.0 | TV-AABC β | 0.047 | 1.05 | 0.213 | — |
| S3k-ped-null: Low heterogeneity (null) | PED | 0.0 | TV-AABC γ | 0.064 | 1.03 | 0.089 | — |
| S3l-bio: High heterogeneity | BIO | 0.5 | Standard countdown | 0.909 | 1.02 | 0.007 | 0.1% |
| S3l-bio: High heterogeneity | BIO | 0.5 | TV-BC | 1.000 | 2.47 | 0.416 | 100.0% |
| S3l-bio: High heterogeneity | BIO | 0.5 | TV-AABC β | 0.993 | 2.08 | 0.360 | 100.0% |
| S3l-bio: High heterogeneity | BIO | 0.5 | TV-AABC γ | 0.990 | 0.65 | 0.066 | 100.0% |
| S3l-bio-null: High heterogeneity (null) | BIO | 0.0 | Standard countdown | 0.996 | 1.04 | 0.008 | — |
| S3l-bio-null: High heterogeneity (null) | BIO | 0.0 | TV-BC | 0.054 | 1.01 | 0.161 | — |

| | | | | | | | |
|---|---|---|---|---|---|---|---|
| S3l-bio-null: High heterogeneity (null) | BIO | 0.0 | TV-AABC β | 0.051 | 1.01 | 0.165 | — |
| S3l-bio-null: High heterogeneity (null) | BIO | 0.0 | TV-AABC γ | 0.037 | 1.01 | 0.110 | — |
| S3l-ped: High heterogeneity | PED | 0.5 | Standard countdown | 1.000 | 1.05 | 0.007 | 0.0% |
| S3l-ped: High heterogeneity | PED | 0.5 | TV-BC | 1.000 | 2.49 | 0.365 | 100.0% |
| S3l-ped: High heterogeneity | PED | 0.5 | TV-AABC β | 1.000 | 2.19 | 0.334 | 100.0% |
| S3l-ped: High heterogeneity | PED | 0.5 | TV-AABC γ | 0.996 | 0.66 | 0.062 | 100.0% |
| S3l-ped-null: High heterogeneity (null) | PED | 0.0 | Standard countdown | 1.000 | 1.07 | 0.008 | — |
| S3l-ped-null: High heterogeneity (null) | PED | 0.0 | TV-BC | 0.038 | 1.01 | 0.136 | — |
| S3l-ped-null: High heterogeneity | PED | 0.0 | TV-AABC β | 0.041 | 1.01 | 0.140 | — |

| | | | | | | | |
|---|---|---|---|---|---|---|---|
| (null) | | | | | | | |
| S3l-ped-null: High heterogeneity (null) | PED | 0.0 | TV-AABC γ | 0.053 | 1.01 | 0.100 | — |

**Supplementary Table 5.** Study 2 disease progression rate results. Family column: PED = pedagogical scenario; BIO = BIOCARD-resembling scenario.

| **Scenario** | **Family** | **True effect size (β)** | **Analytic approach** | **Rejection rate** | **Mean HR** | **SD HR** | **% Correct direction** |
|---|---|---|---|---|---|---|---|
| S3m-bio: Slow disease | BIO | 0.5 | Standard countdown | 0.271 | 1.02 | 0.011 | 8.0% |
| S3m-bio: Slow disease | BIO | 0.5 | TV-BC | 0.609 | 1.63 | 0.334 | 99.4% |
| S3m-bio: Slow disease | BIO | 0.5 | TV-AABC β | 0.304 | 1.41 | 0.296 | 93.8% |
| S3m-bio: Slow disease | BIO | 0.5 | TV-AABC γ | 0.613 | 0.79 | 0.085 | 98.8% |
| S3m-bio-null: Slow disease | BIO | 0.0 | Standard countdown | 0.637 | 1.03 | 0.012 | — |

| | | | | | | | |
|---|---|---|---|---|---|---|---|
| (null) | | | | | | | |
| S3m-bio-null: Slow disease (null) | BIO | 0.0 | TV-BC | 0.042 | 1.03 | 0.205 | — |
| S3m-bio-null: Slow disease (null) | BIO | 0.0 | TV-AABC β | 0.055 | 1.03 | 0.220 | — |
| S3m-bio-null: Slow disease (null) | BIO | 0.0 | TV-AABC γ | 0.054 | 1.02 | 0.119 | — |
| S3m-ped: Slow disease | PED | 0.5 | Standard countdown | 0.770 | 1.03 | 0.011 | 0.3% |
| S3m-ped: Slow disease | PED | 0.5 | TV-BC | 0.720 | 1.65 | 0.312 | 99.5% |
| S3m-ped: Slow disease | PED | 0.5 | TV-AABC β | 0.452 | 1.47 | 0.291 | 96.9% |
| S3m-ped: Slow disease | PED | 0.5 | TV-AABC γ | 0.600 | 0.80 | 0.084 | 98.4% |
| S3m-ped-null: Slow | PED | 0.0 | Standard countdown | 0.961 | 1.05 | 0.012 | — |

| | | | | | | | |
|---|---|---|---|---|---|---|---|
| disease (null) | | | | | | | |
| S3m-ped-null: Slow disease (null) | PED | 0.0 | TV-BC | 0.046 | 1.02 | 0.187 | — |
| S3m-ped-null: Slow disease (null) | PED | 0.0 | TV-AABC β | 0.045 | 1.03 | 0.197 | — |
| S3m-ped-null: Slow disease (null) | PED | 0.0 | TV-AABC γ | 0.056 | 1.02 | 0.114 | — |
| S3n-bio: Fast disease | BIO | 0.5 | Standard countdown | 0.618 | 1.02 | 0.009 | 1.2% |
| S3n-bio: Fast disease | BIO | 0.5 | TV-BC | 0.828 | 1.61 | 0.260 | 99.5% |
| S3n-bio: Fast disease | BIO | 0.5 | TV-AABC β | 0.545 | 1.42 | 0.233 | 97.7% |
| S3n-bio: Fast disease | BIO | 0.5 | TV-AABC γ | 0.752 | 0.80 | 0.069 | 99.9% |
| S3n-bio-null: Fast | BIO | 0.0 | Standard countdown | 0.907 | 1.03 | 0.009 | — |

| | | | | | | | |
|---|---|---|---|---|---|---|---|
| disease (null) | | | | | | | |
| S3n-bio-null: Fast disease (null) | BIO | 0.0 | TV-BC | 0.048 | 1.01 | 0.158 | — |
| S3n-bio-null: Fast disease (null) | BIO | 0.0 | TV-AABC β | 0.041 | 1.02 | 0.162 | — |
| S3n-bio-null: Fast disease (null) | BIO | 0.0 | TV-AABC γ | 0.049 | 1.02 | 0.091 | — |
| S3n-ped: Fast disease | PED | 0.5 | Standard countdown | 0.999 | 1.04 | 0.008 | 0.0% |
| S3n-ped: Fast disease | PED | 0.5 | TV-BC | 0.955 | 1.60 | 0.211 | 100.0% |
| S3n-ped: Fast disease | PED | 0.5 | TV-AABC β | 0.802 | 1.47 | 0.198 | 99.5% |
| S3n-ped: Fast disease | PED | 0.5 | TV-AABC γ | 0.797 | 0.81 | 0.064 | 99.7% |
| S3n-ped-null: Fast disease (null) | PED | 0.0 | Standard countdown | 1.000 | 1.06 | 0.009 | — |
| S3n-ped- | PED | 0.0 | TV-BC | 0.047 | 1.01 | 0.129 | — |

| | | | | | | | |
|---|---|---|---|---|---|---|---|
| null: Fast disease (null) | | | | | | | |
| S3n-ped-null: Fast disease (null) | PED | 0.0 | TV-AABC β | 0.040 | 1.01 | 0.133 | — |
| S3n-ped-null: Fast disease (null) | PED | 0.0 | TV-AABC γ | 0.058 | 1.01 | 0.081 | — |

**Supplementary Table 6.** Study 2 measurement precision results. Family column: PED = pedagogical scenario; BIO = BIOCARD-resembling scenario.

| **Scenario** | **Family** | **True effect size (β)** | **Analytic approach** | **Rejection rate** | **Mean HR** | **SD HR** | **% Correct direction** |
|---|---|---|---|---|---|---|---|
| S3o-ped: Frequent visits | PED | 0.5 | Standard countdown | 0.938 | 1.04 | 0.009 | 0.0% |
| S3o-ped: Frequent visits | PED | 0.5 | TV-BC | 0.828 | 1.61 | 0.263 | 99.8% |
| S3o-ped: Frequent visits | PED | 0.5 | TV-AABC β | 0.558 | 1.44 | 0.238 | 98.8% |
| S3o-ped: | PED | 0.5 | TV-AABC | 0.746 | 0.79 | 0.073 | 99.4% |

| | | | | | | | |
|---|---|---|---|---|---|---|---|
| Frequent visits | | | γ | | | | |
| S3o-ped-null: Frequent visits (null) | PED | 0.0 | Standard countdown | 0.998 | 1.05 | 0.010 | — |
| S3o-ped-null: Frequent visits (null) | PED | 0.0 | TV-BC | 0.045 | 1.02 | 0.158 | — |
| S3o-ped-null: Frequent visits (null) | PED | 0.0 | TV-AABC β | 0.053 | 1.02 | 0.164 | — |
| S3o-ped-null: Frequent visits (null) | PED | 0.0 | TV-AABC γ | 0.049 | 1.00 | 0.095 | — |
| S3p-ped: Sparse visits | PED | 0.5 | Standard countdown | 0.924 | 1.04 | 0.010 | 0.0% |
| S3p-ped: Sparse visits | PED | 0.5 | TV-BC | 0.852 | 1.62 | 0.251 | 100.0% |
| S3p-ped: Sparse visits | PED | 0.5 | TV-AABC β | 0.607 | 1.46 | 0.229 | 99.4% |

| | | | | | | | |
|---|---|---|---|---|---|---|---|
| S3p-ped: Sparse visits | PED | 0.5 | TV-AABC γ | 0.702 | 0.80 | 0.074 | 99.1% |
| S3p-ped-null: Sparse visits (null) | PED | 0.0 | Standard countdown | 0.997 | 1.05 | 0.011 | — |
| S3p-ped-null: Sparse visits (null) | PED | 0.0 | TV-BC | 0.046 | 1.01 | 0.159 | — |
| S3p-ped-null: Sparse visits (null) | PED | 0.0 | TV-AABC β | 0.043 | 1.02 | 0.164 | — |
| S3p-ped-null: Sparse visits (null) | PED | 0.0 | TV-AABC γ | 0.044 | 1.02 | 0.097 | — |
| S3q-bio: High meas. error | BIO | 0.5 | Standard countdown | 0.505 | 1.02 | 0.010 | 2.4% |
| S3q-bio: High meas. error | BIO | 0.5 | TV-BC | 0.618 | 1.54 | 0.277 | 98.8% |

| | | | | | | | |
|---|---|---|---|---|---|---|---|
| S3q-bio: High meas. error | BIO | 0.5 | TV-AABC β | 0.353 | 1.39 | 0.259 | 95.7% |
| S3q-bio: High meas. error | BIO | 0.5 | TV-AABC γ | 0.453 | 0.84 | 0.082 | 96.2% |
| S3q-bio-null: High meas. error (null) | BIO | 0.0 | Standard countdown | 0.806 | 1.03 | 0.010 | — |
| S3q-bio-null: High meas. error (null) | BIO | 0.0 | TV-BC | 0.054 | 1.03 | 0.191 | — |
| S3q-bio-null: High meas. error (null) | BIO | 0.0 | TV-AABC β | 0.057 | 1.05 | 0.204 | — |
| S3q-bio-null: High meas. error | BIO | 0.0 | TV-AABC γ | 0.061 | 1.05 | 0.108 | — |

| | | | | | | | |
|---|---|---|---|---|---|---|---|
| (null) | | | | | | | |
| S3q-ped: High meas. error | PED | 0.5 | Standard countdown | 0.956 | 1.04 | 0.010 | 0.0% |
| S3q-ped: High meas. error | PED | 0.5 | TV-BC | 0.787 | 1.56 | 0.247 | 100.0% |
| S3q-ped: High meas. error | PED | 0.5 | TV-AABC β | 0.581 | 1.45 | 0.238 | 98.4% |
| S3q-ped: High meas. error | PED | 0.5 | TV-AABC γ | 0.437 | 0.85 | 0.077 | 95.9% |
| S3q-ped-null: High meas. error (null) | PED | 0.0 | Standard countdown | 0.995 | 1.05 | 0.011 | — |
| S3q-ped-null: High meas. error (null) | PED | 0.0 | TV-BC | 0.042 | 1.03 | 0.162 | — |
| S3q-ped-null: | PED | 0.0 | TV-AABC β | 0.046 | 1.04 | 0.170 | — |

| | | | | | | | |
|---|---|---|---|---|---|---|---|
| High meas. error (null) | | | | | | | |
| S3q-ped-null: High meas. error (null) | PED | 0.0 | TV-AABC γ | 0.069 | 1.05 | 0.099 | — |

**Supplementary Table 7.** Study 2 cross-product scenario results. Family column: PED = pedagogical scenario; BIO = BIOCARD-resembling scenario.

| Scenario | Family | True effect size (β) | Analytic approach | Rejection rate | Mean HR | SD HR | % Correct direction |
|---|---|---|---|---|---|---|---|
| S3r-bio: Strong harmful + large n | BIO | 1.0 | Standard countdown | 0.116 | 1.01 | 0.006 | 18.9% |
| S3r-bio: Strong harmful + large n | BIO | 1.0 | TV-BC | 1.000 | 2.48 | 0.343 | 100.0% |
| S3r-bio: | BIO | 1.0 | TV-AABC | 0.996 | 1.85 | 0.253 | 100.0% |

| | | | | | | | |
|---|---|---|---|---|---|---|---|
| Strong harmful + large n | | | β | | | | |
| S3r-bio: Strong harmful + large n | BIO | 1.0 | TV-AABC γ | 1.000 | 0.62 | 0.042 | 100.0% |
| S3r-ped: Strong harmful + large n | PED | 1.0 | Standard countdown | 0.853 | 1.02 | 0.006 | 0.1% |
| S3r-ped: Strong harmful + large n | PED | 1.0 | TV-BC | 1.000 | 2.47 | 0.301 | 100.0% |
| S3r-ped: Strong harmful + large n | PED | 1.0 | TV-AABC β | 1.000 | 1.97 | 0.244 | 100.0% |
| S3r-ped: Strong harmful + large n | PED | 1.0 | TV-AABC γ | 1.000 | 0.63 | 0.040 | 100.0% |
| S3s-bio: Strong protective + large n | BIO | -1.0 | Standard countdown | 1.000 | 1.05 | 0.008 | 100.0% |
| S3s-bio: Strong protective + large n | BIO | -1.0 | TV-BC | 1.000 | 0.45 | 0.054 | 100.0% |

| | | | | | | | |
|---|---|---|---|---|---|---|---|
| S3s-bio: Strong protective + large n | BIO | -1.0 | TV-AABC β | 0.997 | 0.53 | 0.071 | 100.0% |
| S3s-bio: Strong protective + large n | BIO | -1.0 | TV-AABC γ | 1.000 | 1.52 | 0.126 | 100.0% |
| S3s-ped: Strong harmful + very large n | PED | 1.0 | Standard countdown | 0.992 | 1.02 | 0.004 | 0.0% |
| S3s-ped: Strong harmful + very large n | PED | 1.0 | TV-BC | 1.000 | 2.48 | 0.207 | 100.0% |
| S3s-ped: Strong harmful + very large n | PED | 1.0 | TV-AABC β | 1.000 | 1.98 | 0.167 | 100.0% |
| S3s-ped: Strong harmful + very large n | PED | 1.0 | TV-AABC γ | 1.000 | 0.63 | 0.029 | 100.0% |
| S3t-ped: Strong protective | PED | -1.0 | Standard countdown | 1.000 | 1.08 | 0.008 | 100.0% |

| | | | | | | | |
|---|---|---|---|---|---|---|---|
| + large n | | | | | | | |
| S3t-ped: Strong protective + large n | PED | -1.0 | TV-BC | 1.000 | 0.44 | 0.045 | 100.0% |
| S3t-ped: Strong protective + large n | PED | -1.0 | TV-AABC β | 1.000 | 0.49 | 0.054 | 100.0% |
| S3t-ped: Strong protective + large n | PED | -1.0 | TV-AABC γ | 1.000 | 1.53 | 0.114 | 100.0% |
| S3u-ped: Strong harmful + high het | PED | 1.0 | Standard countdown | 1.000 | 1.04 | 0.007 | 0.0% |
| S3u-ped: Strong harmful + high het | PED | 1.0 | TV-BC | 1.000 | 5.21 | 0.896 | 100.0% |
| S3u-ped: Strong harmful + high het | PED | 1.0 | TV-AABC β | 1.000 | 4.18 | 0.753 | 100.0% |
| S3u-ped: Strong harmful + high het | PED | 1.0 | TV-AABC γ | 1.000 | 0.45 | 0.043 | 100.0% |
| S3v-ped: Strong | PED | -1.0 | Standard countdown | 1.000 | 1.10 | 0.011 | 100.0% |

| | | | | | | | |
|---|---|---|---|---|---|---|---|
| protective + high het | | | | | | | |
| S3v-ped: Strong protective + high het | PED | -1.0 | TV-BC | 1.000 | 0.22 | 0.033 | 100.0% |
| S3v-ped: Strong protective + high het | PED | -1.0 | TV-AABC β | 1.000 | 0.23 | 0.037 | 100.0% |
| S3v-ped: Strong protective + high het | PED | -1.0 | TV-AABC γ | 1.000 | 2.01 | 0.257 | 100.0% |
| S3w-ped: Strong harmful + low het | PED | 1.0 | Standard countdown | 0.059 | 1.01 | 0.014 | 30.6% |
| S3w-ped: Strong harmful + low het | PED | 1.0 | TV-BC | 0.603 | 1.61 | 0.331 | 99.2% |
| S3w-ped: Strong harmful + low het | PED | 1.0 | TV-AABC β | 0.309 | 1.40 | 0.288 | 93.9% |
| S3w-ped: | PED | 1.0 | TV-AABC | 0.704 | 0.81 | 0.069 | 99.7% |

| | | | | | | | |
|---|---|---|---|---|---|---|---|
| Strong harmful + low het | | | γ | | | | |
| S3x-ped: Strong harmful + fast disease | PED | 1.0 | Standard countdown | 0.911 | 1.03 | 0.008 | 0.0% |
| S3x-ped: Strong harmful + fast disease | PED | 1.0 | TV-BC | 1.000 | 2.43 | 0.335 | 100.0% |
| S3x-ped: Strong harmful + fast disease | PED | 1.0 | TV-AABC β | 0.999 | 2.03 | 0.280 | 100.0% |
| S3x-ped: Strong harmful + fast disease | PED | 1.0 | TV-AABC γ | 1.000 | 0.65 | 0.053 | 100.0% |
| S3y-ped: Strong harmful + high error | PED | 1.0 | Standard countdown | 0.663 | 1.02 | 0.009 | 0.9% |
| S3y-ped: Strong harmful + high error | PED | 1.0 | TV-BC | 1.000 | 2.24 | 0.356 | 100.0% |

| | | | | | | | |
|---|---|---|---|---|---|---|---|
| S3y-ped: Strong harmful + high error | PED | 1.0 | TV-AABC β | 0.974 | 1.89 | 0.301 | 100.0% |
| S3y-ped: Strong harmful + high error | PED | 1.0 | TV-AABC γ | 0.959 | 0.71 | 0.064 | 100.0% |
| S3z-ped: Max power scenario | PED | 1.0 | Standard countdown | 1.000 | 1.03 | 0.004 | 0.0% |
| S3z-ped: Max power scenario | PED | 1.0 | TV-BC | 1.000 | 2.40 | 0.163 | 100.0% |
| S3z-ped: Max power scenario | PED | 1.0 | TV-AABC β | 1.000 | 2.00 | 0.139 | 100.0% |
| S3z-ped: Max power scenario | PED | 1.0 | TV-AABC γ | 1.000 | 0.64 | 0.026 | 100.0% |

**Supplementary Table 8. Full model coefficients for all Cox proportional hazards models. Each block shows one biomarker and analysis method. Covariates: sex, education (standardized), and APOE ε4 carrier status.**

| **Parameter** | **HR (95% CI; P)** |
|---|---|
| **BIOCARD – CSF Aβ42/Aβ40 — Standard countdown (n = 114; 54 clinical onsets)** | |
| Sex (female) | 0.75 (0.39–1.43); P = 0.39 |
| Education (z) | 0.99 (0.73–1.33); P = 0.92 |
| APOEε4 | 0.80 (0.45–1.43); P = 0.45 |
| AABC (z) | 2.08 (1.44–3.01); P < 0.001 |
| **BIOCARD – CSF Aβ42/Aβ40 — TV-BC (n = 238; 83 clinical onsets)** | |
| Sex (female) | 0.94 (0.58–1.54); P = 0.81 |
| Education (z) | 0.98 (0.77–1.23); P = 0.83 |
| APOEε4 | 1.06 (0.65–1.73); P = 0.80 |
| Biomarker positivity | 3.02 (1.80–5.05); P < 0.001 |
| **BIOCARD – CSF Aβ42/Aβ40 — TV-AABC (n = 238; 83 clinical onsets)** | |
| Sex (female) | 0.93 (0.57–1.53); P = 0.79 |
| Education (z) | 0.97 (0.77–1.23); P = 0.82 |
| APOEε4 | 0.94 (0.56–1.56); P = 0.80 |
| Biomarker positivity | 3.25 (1.93–5.48); P < 0.001 |
| Biomarker positivity × AABC | 0.71 (0.51–0.99); P < 0.05 |
| **BIOCARD – CSF p-tau181 — Standard countdown (n = 142; 56 clinical onsets)** | |
| Sex (female) | 0.83 (0.45–1.55); P = 0.57 |
| Education (z) | 1.01 (0.74–1.37); P = 0.97 |
| APOEε4 | 1.19 (0.67–2.10); P = 0.55 |
| AABC (z) | 2.96 (1.99–4.39); P < 0.001 |
| **BIOCARD – CSF p-tau181 — TV-BC (n = 238; 83 clinical onsets)** | |
| Sex (female) | 1.04 (0.64–1.67); P = 0.88 |
| Education (z) | 0.96 (0.76–1.21); P = 0.75 |

| | |
|---|---|
| APOEε4 | 1.52 (0.95–2.43); P = 0.08 |
| Biomarker positivity | 1.52 (0.91–2.54); P = 0.11 |
| **BIOCARD – CSF p-tau181 — TV-AABC (n = 238; 83 clinical onsets)** | |
| Sex (female) | 0.95 (0.58–1.54); P = 0.83 |
| Education (z) | 0.96 (0.76–1.22); P = 0.75 |
| APOEε4 | 1.24 (0.77–2.01); P = 0.38 |
| Biomarker positivity | 1.45 (0.87–2.40); P = 0.15 |
| Biomarker positivity × AABC | 0.58 (0.42–0.78); P < 0.001 |
| **BIOCARD – plasma p-tau181 — Standard countdown (n = 95; 29 clinical onsets)** | |
| Sex (female) | 1.59 (0.65–3.87); P = 0.31 |
| Education (z) | 0.98 (0.64–1.48); P = 0.91 |
| APOEε4 | 1.88 (0.86–4.11); P = 0.11 |
| AABC (z) | 2.34 (1.39–3.94); P < 0.01 |
| **BIOCARD – plasma p-tau181 — TV-BC (n = 200; 37 clinical onsets)** | |
| Sex (female) | 1.39 (0.67–2.91); P = 0.38 |
| Education (z) | 0.91 (0.66–1.25); P = 0.55 |
| APOEε4 | 2.23 (1.12–4.43); P < 0.05 |
| Biomarker positivity | 3.28 (1.42–7.61); P < 0.01 |
| **BIOCARD – plasma p-tau181 — TV-AABC (n = 200; 37 clinical onsets)** | |
| Sex (female) | 1.49 (0.71–3.14); P = 0.29 |
| Education (z) | 0.91 (0.66–1.26); P = 0.56 |
| APOEε4 | 2.37 (1.19–4.74); P < 0.05 |
| Biomarker positivity | 2.74 (1.18–6.36); P < 0.05 |
| Biomarker positivity × AABC | 0.51 (0.35–0.74); P < 0.001 |
| **ADNI – amyloid PET — Standard countdown (n = 236; 66 clinical onsets)** | |

| | |
|---|---|
| Sex (female) | 0.82 (0.47–1.42); P = 0.47 |
| Education (z) | 0.90 (0.71–1.14); P = 0.39 |
| APOEε4 | 1.31 (0.80–2.17); P = 0.28 |
| AABC (z) | 3.34 (2.41–4.61); P < 0.001 |
| **ADNI – amyloid PET — TV-BC (n = 575; 106 clinical onsets)** | |
| Sex (female) | 0.66 (0.44–0.99); P < 0.05 |
| Education (z) | 0.87 (0.72–1.05); P = 0.14 |
| APOEε4 | 1.07 (0.70–1.61); P = 0.76 |
| Biomarker positivity | 2.77 (1.84–4.17); P < 0.001 |
| **ADNI – amyloid PET — TV-AABC (n = 575; 106 clinical onsets)** | |
| Sex (female) | 0.64 (0.43–0.97); P < 0.05 |
| Education (z) | 0.87 (0.72–1.05); P = 0.13 |
| APOEε4 | 1.05 (0.69–1.59); P = 0.82 |
| Biomarker positivity | 2.89 (1.91–4.37); P < 0.001 |
| Biomarker positivity × AABC | 0.87 (0.64–1.18); P = 0.36 |
| **ADNI – plasma p-tau217 — Standard countdown (n = 122; 48 clinical onsets)** | |
| Sex (female) | 1.43 (0.73–2.79); P = 0.30 |
| Education (z) | 1.03 (0.75–1.43); P = 0.84 |
| APOEε4 | 1.29 (0.70–2.38); P = 0.41 |
| AABC (z) | 1.94 (1.34–2.80); P < 0.001 |
| **ADNI – plasma p-tau217 — TV-BC (n = 801; 162 clinical onsets)** | |
| Sex (female) | 0.68 (0.50–0.94); P < 0.05 |
| Education (z) | 0.84 (0.72–0.97); P < 0.05 |
| APOEε4 | 1.02 (0.73–1.41); P = 0.93 |
| Biomarker positivity | 2.60 (1.83–3.69); P < 0.001 |
| **ADNI – plasma p-tau217 — TV-AABC** | |

| | |
|---|---|
| **(n = 801; 162 clinical onsets)** | |
| Sex (female) | 0.67 (0.49–0.92); $P < 0.05$ |
| Education (z) | 0.82 (0.71–0.96); $P < 0.05$ |
| APOEε4 | 1.00 (0.72–1.38); $P = 0.98$ |
| Biomarker positivity | 2.89 (2.05–4.08); $P < 0.001$ |
| Biomarker positivity × AABC | 0.46 (0.32–0.66); $P < 0.001$ |
